# Explainable AI to predict a complex multifactorial outcome, childhood obesity: Application to clinical epidemiology

**DOI:** 10.1101/2025.06.21.25330041

**Authors:** Fuling Chen, Phillip E Melton, Kevin Vinsen, Trevor Mori, Lawrence Beilin, Rae-Chi Huang

**Affiliations:** International Centre for Radio Astronomy Research, University of Western Australia, 35 Stirling Hwy, Perth, 6009, WA, Australia; Menzies Institute for Medical Research, University of Tasmania, Churchill Ave, Hobart, 7005, TAS, Australia; Medical School, University of Western Australia, 35 Stirling Hwy, Perth, 6009, WA, Australia; Nutrition & Health Innovation Research Institute, Edith Cowan University, 2 Bradford Street, Perth, 6050, EA, Australia

**Keywords:** Machine learning, Explainable AI, Obesity, Epidemiology, Body Mass Index

## Abstract

**Background:** Childhood obesity, driven by genetic and epidemiological factors, poses significant health risks, yet traditional machine learning models lack interpretability for clinical use.

**Objective:** This study aims to apply Kolmogorov-Arnold Networks (KAN), an explainable machine learning model, to predict body mass index (BMI) at age 8 as an indicator of obesity risk and to develop a publicly accessible prediction tool.

**Methods:** We utilized the Raine Study Gen2 cohort (n=2,868) to train KAN and traditional models (such as Random Forest, Gradient Boosting, Lasso, and Multi-Layer Perceptron) using perinatal, early-life, and polygenic risk score (PGS) data collected before age 5. Feature importance was analyzed across all the models. A publicly accessible online calculator was developed for practical use.

**Results:** KAN achieved an R^2^ of 0.81, outperforming traditional models. Key predictors included Year 5 BMI z-score, mid-arm circumference, occupation of mother, and PGS. The online calculator supports predictions without PGS, maintaining an R^2^ of 0.81.

**Conclusions:** KAN’s transparent formulas enhance interpretability, offering a practical approach to predicting childhood obesity. The freely accessible tool enables clinicians to implement personalized prevention strategies, advancing precision medicine.

**Graphical abstract:** KAN model predicts childhood obesity (BMI at age 8), showcasing key features, top performance, and accurate formularised results with epidemiological and genetic factors. Online calculator is available at https://bmi-y8-calc.onrender.com/.

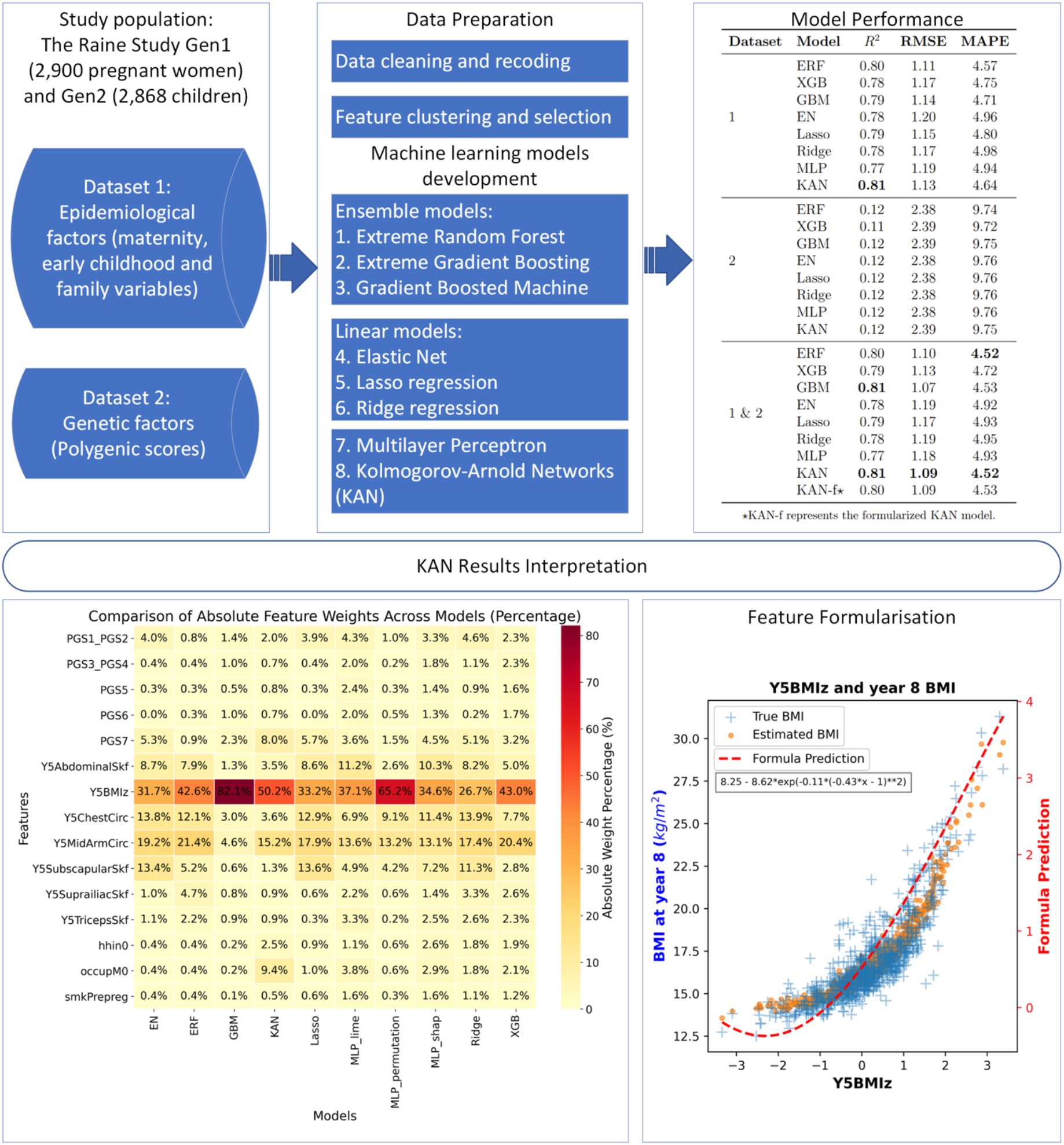

## 1 Introduction

During the past decade, artificial intelligence, particularly machine learning (ML) models, has played a transformative role in predicting complex diseases. Obesity, a multifaceted disease driven by both environmental and genetic factors, has been widely studied using body mass index (BMI) as a key surrogate measure of health risk at the population level ^1^. Reviews focusing on ML applications in this field ^2,3^ highlight that numerous models have been developed to predict BMI. The popularly used models include Lasso ^4^, Logistic Regression (LR) ^5^, Random Forests (RF) ^6^, Support Vector Machines (SVM) ^7^, Extreme Gradient Boosting (XGB) ^8^, Artificial Neural Networks (ANN) ^9,10^, and Long Short-Term Memory network (LSTM) ^11^.

The systematic reviews by Colmenarejo ^2^ and Safaei et al. ^3^ underscore the advantages of ML models, particularly their superior predictive accuracy, which is often achieved through ensemble methods and deep learning models that frequently outperform traditional statistical techniques. ML models also capture complexity and nonlinear relationships among obesity-related variables. Nonetheless, significant challenges remain. Many ML models, especially deep learning, are criticized for their lack of interpretability, often functioning as “black boxes”. To address this, SHapley Additive exPlanations (SHAP), Local Interpretable Model-agnostic Explanations (LIME), and permutation importance are widely used to interpret Multi-Layer Perceptrons (MLPs) ^12–14^. These methods provide variable contributions; however, they may yield inconsistent feature importance values and struggle to fully clarify decision-making in deep models, raising concerns about reliability ^15^.

Kolmogorov-Arnold Networks (KAN) represent a recently developed deep learning model ^16^. By eliminating linear weights entirely and replacing them with univariate functions parametrized as splines, this innovative architecture is believed to enhance the model’s accuracy compared to the MLP, claiming superior performance. It requires smaller and shallower architectures to achieve faster neural scaling laws, thus improving computational efficiency.

Increased interpretability is highly valued in scenarios that require transparency and explanation ^17,18^, especially in clinical scenarios. When clinical decisions are being made, potentially affecting an individual’s course of care, understanding of the logic is reassuring, if not mandatory, rather than devolving decision making to a “black box”. The KAN model’s intrinsic architecture delivers explicit mathematical formulas for decision-making, enhancing its translational impact over other deep learning models by enabling users to make predictions using only a scientific calculator for practical clinical applications. Unlike traditional machine learning models, which often require substantial computational resources such as GPUs, CPUs, or cloud services, KAN’s approach overcomes these barriers, significantly improving its utility in real-world clinical settings.

In this study, we aimed to demonstrate the application of a KAN to predict childhood obesity, a complex condition influenced by both biological and environmental factors, by predicting BMI at age 8, elucidating the model’s decision-making process through transparent mathematical formulas, and implementing a publicly accessible, user-friendly tool for general clinical use. The transition from childhood to adulthood represents a critical period for obesity prevention. Longitudinal studies indicate that approximately 55% children with obesity remain with excess weight in adolescence, 80% of adolescents continue to be with obesity as adults, and 70% maintain obesity over age 30 ^19^ These findings underscore the long-term health implications of childhood obesity and highlight the importance of early intervention.

This study makes contributions to the field of clinical epidemiology by pioneering the application of KAN to predict childhood obesity at age 8, achieving a superior R^2^ of 0.81 using the Raine Study Gen2 cohort. KAN’s transparent mathematical formulas elucidate key predictors, including BMI z-scores, mid-arm circumference at the age of 5, maternal occupation, and genetic factor, providing actionable insights for clinicians (see Table 1). A publicly accessible Year 8 BMI Prediction Calculator, hosted on Render and GitHub, enables clinicians, researchers, and parents caring for young children to make interpretable predictions without requiring genetic data, thereby supporting early interventions for obesity.

**Table 1.** Statement of significance.

|  |
| --- |
| **Problem** |
| Childhood obesity is a complex, multifactorial condition driven by genetic, environmental, and early-life factors, making accurate prediction with reasoning challenging with traditional machine learning models due to their limited interpretability. |
| **What is Already Known** |
| Traditional machine learning models have been applied to predict obesity using BMI as a surrogate, showing high accuracy but suffering from "black box" opacity. Kolmogorov-Arnold Networks (KAN) provide intrinsic interpretability as an alternative, but no prior studies have applied them to childhood obesity prediction. |
| **What This Paper Adds** |
| This study applied an explainable AI model (KAN) to predict BMI at age 8 in the Raine Study cohort, achieving a $R^2$ of 0.81 with transparent mathematical formulas that highlight key predictors like year-5 BMI z-score, mid-arm circumference, maternal occupation, and polygenic scores; it also introduces a publicly accessible online calculator for predictions without genetic data. |
| **Who would benefit** |
| Clinicians seeking interpretable tools for early obesity risk assessment; researchers in precision medicine exploring explainable AI; and parents or caregivers. |

## 2 Methods

### 2.1 Study Population

This study utilized data from the Raine Study Gen2^1^. The Raine Study is a large, well-established longitudinal cohort designed to track health and developmental outcomes from pregnancy through to adulthood ^20^. The Raine Study initially recruited 2,900 pregnant women (Generation 1, Gen1) and followed 2,868 of their children (Generation 2, Gen2). Recruitment was primarily conducted at King Edward Memorial Hospital in Perth, Western Australia, making the study population representative of the regional population. Dataset 1 collection time points were at 18, 24, 34 weeks’ gestation during pregnancy, at birth, and at 1 and 5 years of age. The outcome measure data collection occurred at the age of 8 years.

### 2.2 Dataset 1: Epidemiological Factors

In this study, Dataset 1 was created to explore the influence of maternal, early childhood, and family factors on BMI development over time.

This dataset initially included 201 raw variables (Step 1) spanning various domains, such as maternal characteristics (eg, weight gain and smoking status during pregnancy), early childhood physical measures (eg, weight and height at birth, year 1 and year 5, breastfeeding duration and childcare status), and family demographics (eg, parental education, occupation, and household income). The details of the variables are provided in the Supplementary Material.

The processing of dataset 1 involved obtaining the raw data, excluding the anomalies, data cleansing, encoding variables, feature selection and clustering. Figure 1 depicts the data flow and the corresponding sample size and number of variables at each step.

**Figure 1.**
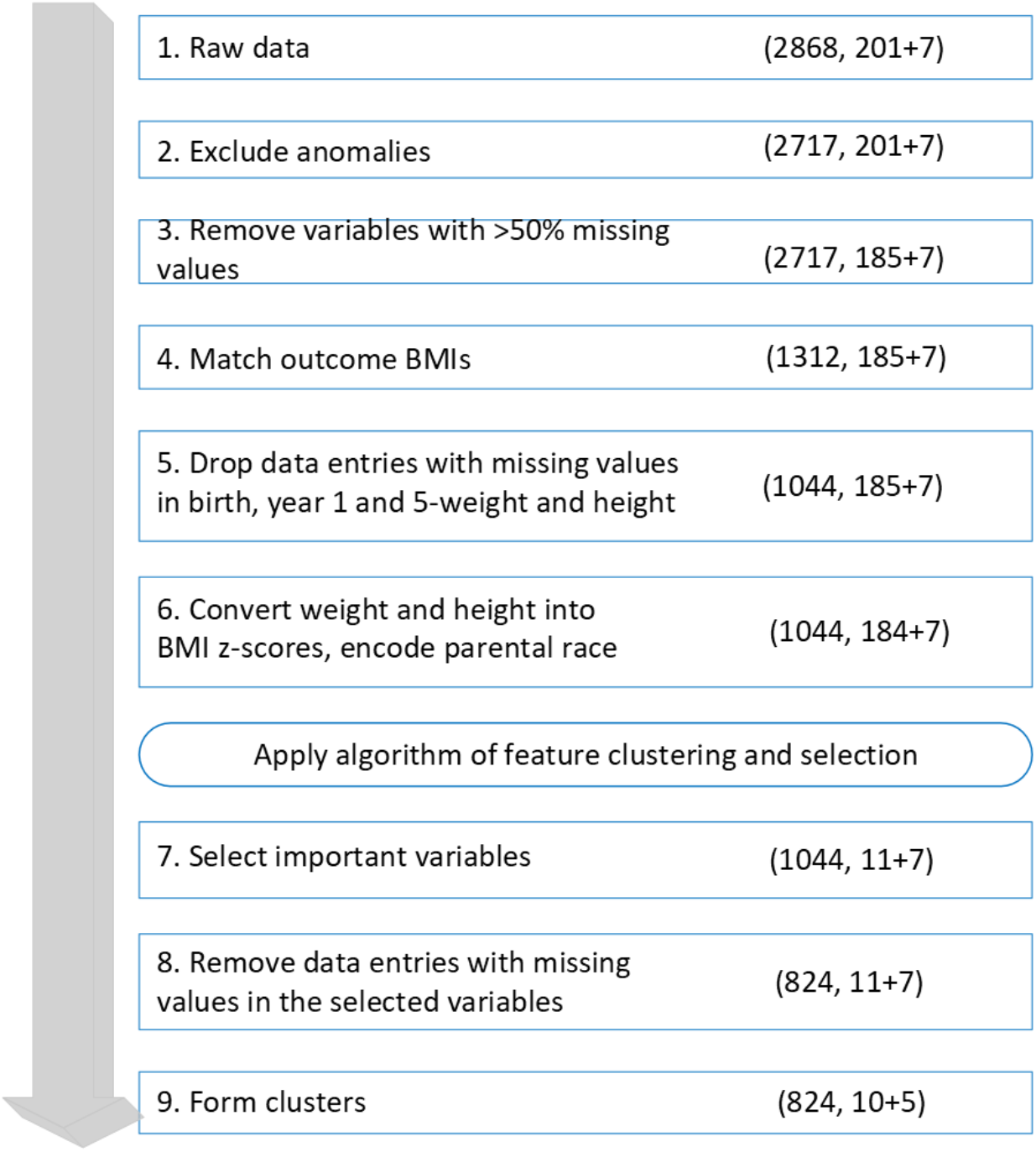
Datasets preprocessing workflow. The form of (i, j+k) represents the number of samples i, the number of dataset 1 variables j, and the number of polygenic scores in dataset 2 (k).

Data preparation involves rigorous cleaning and filtering stages to enhance data quality and minimize bias. We began by excluding cases with significant congenital anomalies or multiple births to ensure a focus on children with typical developmental trajectories, which helps in drawing more generalizable conclusions regarding BMI development (Step 2).

Subsequent filtering focused on addressing the high level of missing data among certain variables, a common challenge in cohort studies. To address this, we removed variables where more than 50% of the values were missing (Step 3), which aligns with best practices for ensuring sufficient data density ^22^. Then, we removed cases without target BMI data (Step 4) and retained cases with key BMI-related data, such as weight and height at birth, and ages at 1 and 5 years (Step 5). These physical measurements were essential for calculating BMI z-scores, which provided a standardized approach for assessing changes in BMI relative to age and gender. To ensure accuracy across different age groups, we used the CDC Extended BMI-for-age growth charts for very high BMI values at age five, given the chart’s improved reliability in capturing elevated BMIs above the 95th percentile ^23^. For younger age points (birth and year one), we applied the World Health Organization’s growth standards, which are widely recognized as appropriate for early developmental stages ^24^ (Step 6).

Dataset 1 also included categorical variables for parental race and ethnicity, represented using one-hot encoding (Step 6), which converts each category into a binary vector for compatibility with machine learning algorithms. This encoding captured demographic diversity with five categories (Caucasian, Aboriginal, Polynesian, Asian, and Other), enabling a more nuanced understanding of BMI development across ethnic backgrounds, as genetic and cultural factors may influence childhood BMI trajectories ^25,26^.

Despite these steps, several variables were found to be strongly correlated, and some variables continued to exhibit missing data. To form independent variables and maximize data retention while minimizing redundancy, we applied a correlation-based clustering algorithm, which groups highly correlated variables, thereby facilitating the identification of core features that contribute independently to BMI prediction. Clustering was followed by importance-based variable selection, which enabled us to prioritize variables with the most significant predictive value for BMI outcomes (see Appendix 1 for more details). This combination of clustering and selection has been shown to enhance model performance by retaining relevant variables while reducing overfitting due to extraneous data ^27^. We obtained the cluster patterns and selected the inclusive variables (Step 7). Finally, we excluded cases without data on selected key variables (Step 8) and formed clusters (Step 9), resulting in a dataset optimized for our subsequent analyses.

After preprocessing, the final dataset includes 824 samples and 10 variables. The baseline characteristics and descriptive statistics of the selected variables are shown in Table 2. Among these variables, smoking before pregnancy (*smkPrepreg*) and cigarettes per day before pregnancy (*cigPrepreg*) were combined into a single factor, hereafter referred to as *smkPrepreg*.

**Table 2.**
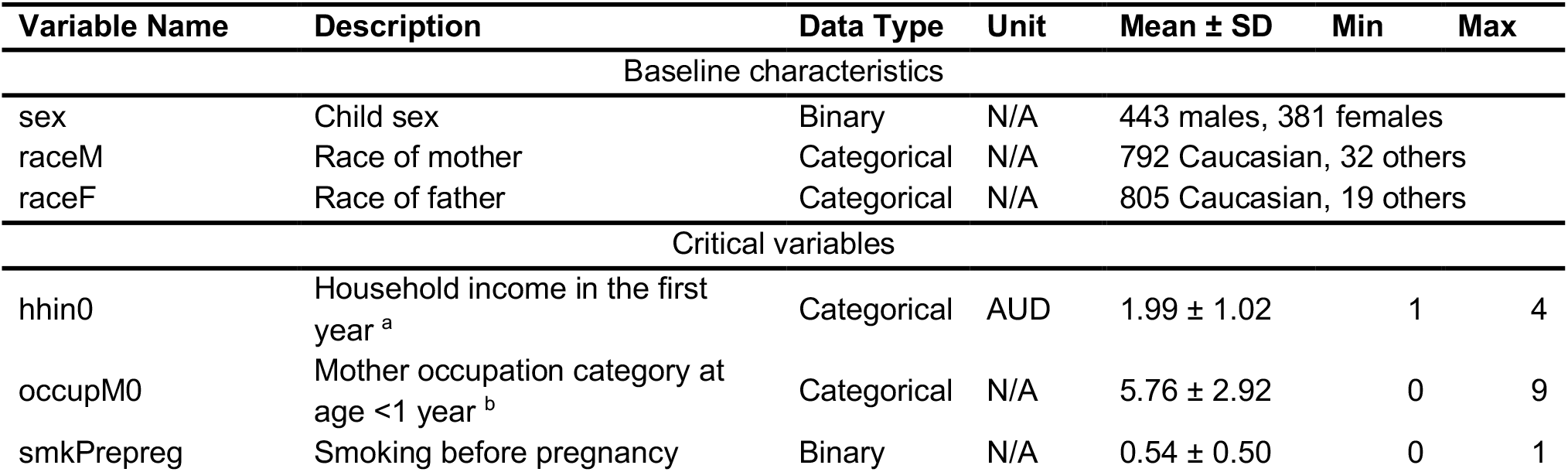
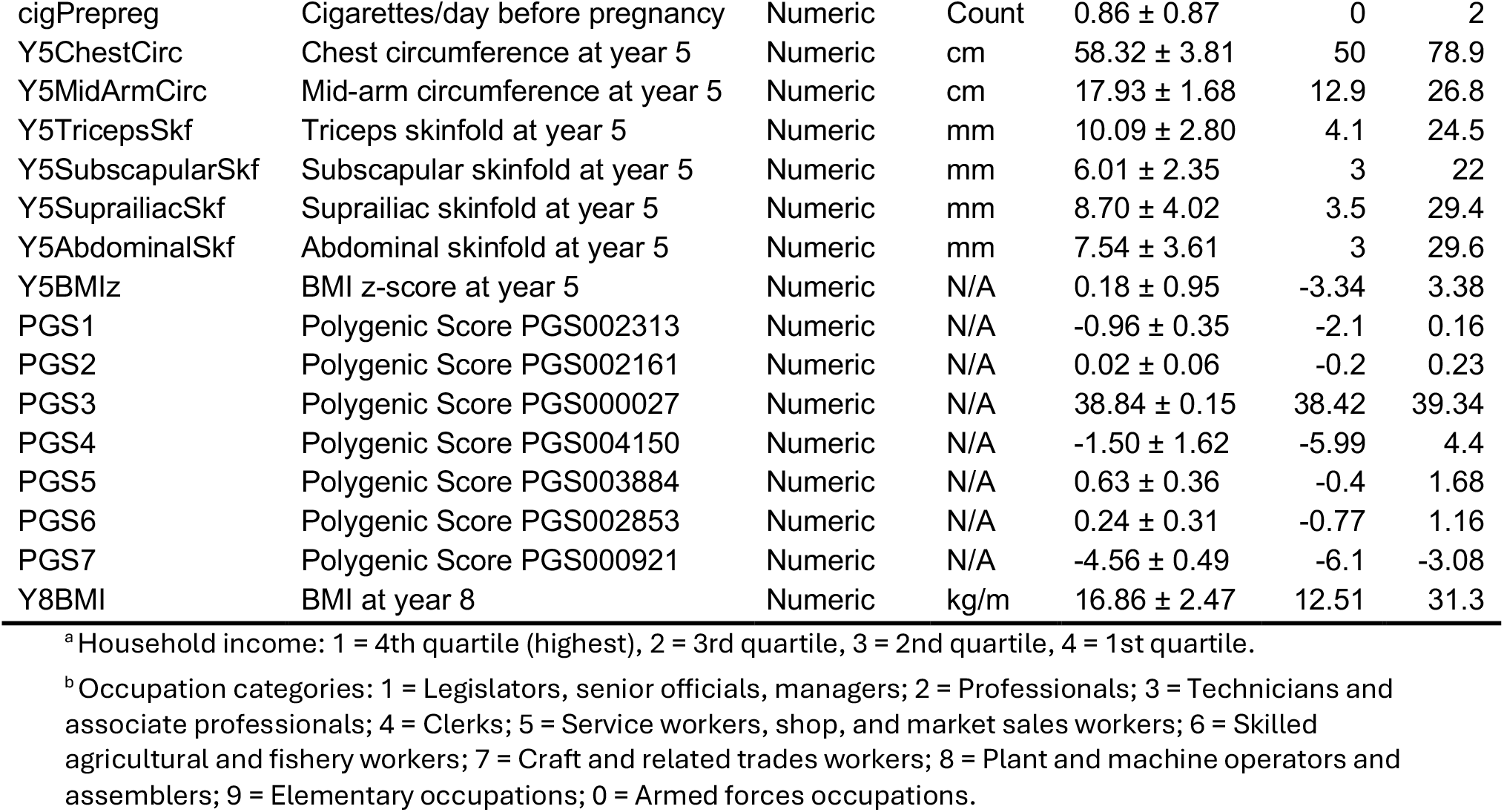
Baseline characteristics and descriptive statistics of critical variables used in the study.

| Variable Name | Description | Data Type | Unit | Mean ± SD | Min | Max |
| --- | --- | --- | --- | --- | --- | --- |
| Baseline characteristics |  |  |  |  |  |  |
| sex | Child sex | Binary | N/A | 443 males, 381 females |  |  |
| raceM | Race of mother | Categorical | N/A | 792 Caucasian, 32 others |  |  |
| raceF | Race of father | Categorical | N/A | 805 Caucasian, 19 others |  |  |
| Critical variables |  |  |  |  |  |  |
| hhin0 | Household income in the first year <sup>a</sup> | Categorical | AUD | 1.99 ± 1.02 | 1 | 4 |
| occupM0 | Mother occupation category at age <1 year <sup>b</sup> | Categorical | N/A | 5.76 ± 2.92 | 0 | 9 |
| smkPrepreg | Smoking before pregnancy | Binary | N/A | 0.54 ± 0.50 | 0 | 1 |
| cigPrepreg | Cigarettes/day before pregnancy | Numeric | Count | 0.86 ± 0.87 | 0 | 2 |
| Y5ChestCirc | Chest circumference at year 5 | Numeric | cm | 58.32 ± 3.81 | 50 | 78.9 |
| Y5MidArmCirc | Mid-arm circumference at year 5 | Numeric | cm | 17.93 ± 1.68 | 12.9 | 26.8 |
| Y5TricepsSkf | Triceps skinfold at year 5 | Numeric | mm | 10.09 ± 2.80 | 4.1 | 24.5 |
| Y5SubscapularSkf | Subscapular skinfold at year 5 | Numeric | mm | 6.01 ± 2.35 | 3 | 22 |
| Y5SuprailiacSkf | Suprailiac skinfold at year 5 | Numeric | mm | 8.70 ± 4.02 | 3.5 | 29.4 |
| Y5AbdominalSkf | Abdominal skinfold at year 5 | Numeric | mm | 7.54 ± 3.61 | 3 | 29.6 |
| Y5BMIz | BMI z-score at year 5 | Numeric | N/A | 0.18 ± 0.95 | -3.34 | 3.38 |
| PGS1 | Polygenic Score PGS002313 | Numeric | N/A | -0.96 ± 0.35 | -2.1 | 0.16 |
| PGS2 | Polygenic Score PGS002161 | Numeric | N/A | 0.02 ± 0.06 | -0.2 | 0.23 |
| PGS3 | Polygenic Score PGS000027 | Numeric | N/A | 38.84 ± 0.15 | 38.42 | 39.34 |
| PGS4 | Polygenic Score PGS004150 | Numeric | N/A | -1.50 ± 1.62 | -5.99 | 4.4 |
| PGS5 | Polygenic Score PGS003884 | Numeric | N/A | 0.63 ± 0.36 | -0.4 | 1.68 |
| PGS6 | Polygenic Score PGS002853 | Numeric | N/A | 0.24 ± 0.31 | -0.77 | 1.16 |
| PGS7 | Polygenic Score PGS000921 | Numeric | N/A | -4.56 ± 0.49 | -6.1 | -3.08 |
| Y8BMI | BMI at year 8 | Numeric | kg/m | 16.86 ± 2.47 | 12.51 | 31.3 |

<sup>a</sup>Household income: 1 = 4th quartile (highest), 2 = 3rd quartile, 3 = 2nd quartile, 4 = 1st quartile.
<sup>b</sup>Occupation categories: 1 = Legislators, senior officials, managers; 2 = Professionals; 3 = Technicians and associate professionals; 4 = Clerks; 5 = Service workers, shop, and market sales workers; 6 = Skilled agricultural and fishery workers; 7 = Craft and related trades workers; 8 = Plant and machine operators and assemblers; 9 = Elementary occupations; 0 = Armed forces occupations.

### 2.3 Dataset 2: Genetic Factors

In addition to Dataset 1, we incorporated seven published BMI polygenic scores (PGS), as outlined in Table 3. These scores were generated using the pgsc_calc pipeline ^28^, which computes PGS by matching genetic variants from scoring files published in the Polygenic Score Catalog with those in the target dataset.

**Table 3.** Summary of polygenic scores (PGS) used in this study. “NR” indicates not reported.

| PGS ID | Reference | PGS Pub. ID | No. Variants | Development Method | Score Development |  |  |
| --- | --- | --- | --- | --- | --- | --- | --- |
|  |  |  |  |  | No. Sample | Ancestry | Cohort |
| PGS1 | <sup>29</sup> | PGP000332 | 1109311 | BOLT-LMM | NR | NR | NR |
| PGS2 | <sup>30</sup> | PGP000263 | 990022 | LDpred2 (bigsnpr) | 391124 | European | UKB |
| PGS3 | <sup>31</sup> | PGP000017 | 2100302 | LDpred | 119951 | European | UKB |
| PGS4 | <sup>32</sup> | PGP000517 | 1039042 | UKBB-EUR.MultiPRS.CV | 359913 | European | UKB |
| PGS5 | <sup>33</sup> | PGP000501 | 1067771 | PRS-CSx | NR | NR | NR |
| PGS6 | <sup>34</sup> | PGP000393 | 7446664 | DBSLMM | 23381 | European | MGI |
| PGS7 | <sup>35</sup> | PGP000243 | 1947711 | LDpred | 1000 | European<br>(Danish) | Inter9<br>9 |

Preprocessing was performed using Dataset 1, Dataset 2, and a combination of both datasets for predicting the outcome of BMI at 17 years old. When merging Dataset 1 and Dataset 2, the seven PGS were treated as extra variables and integrated with those in Dataset 1. After clustering, these seven PGS were grouped into five independent variables, with PGS1 and PGS2 forming one cluster, and PGS3 and PGS4 forming another cluster. The combined data workflow is illustrated in Figure 1, with the final number of clusters being 15. The metadata of the seven PGS are shown in Table 3.

### 2.4 Outcome Measures

For this study, weight and height measurements were collected at the age of 8 years. BMI was calculated by weight divided by height squared, serving as the primary outcome variable. The year 8 BMI is presented in Table 2.

### 2.5 Ethical Considerations

This study, utilizing de-identified data from the Raine Study Gen2 cohort, received an exemption from ethics review by the University of Western Australia’s Human Research Ethics Office (Ref: 2025/ET000396, April 28, 2025), as no new data from human subjects were collected. Informed consent was obtained from participants’ guardians during the original Raine Study, adhering to institutional protocols. Data privacy and confidentiality were ensured through de-identification and secure storage, compliant with Australian data protection guidelines. No compensation was provided to participants, as the study relied on existing data.

### 2.6 Machine Learning Models

This study used seven widely used machine learning models and the KAN ^16^ to analyze the data. The conventional models included three ensemble methods (Extreme Random Forest (ERF) ^36^, Extreme Gradient Boosting (XGB) ^37^, and Gradient Boosted Machine (GBM) ^38^), three linear models (Elastic Net (EN) ^39^, Lasso ^40^, and Ridge ^41^ regression), and Multi-Layer Perceptron (MLP) ^42^. These models were selected for their strong performance in regression tasks and high interpretability, making them well-suited for understanding the relationships within the data. The conventional models were implemented using the Python package scikit-learn (version 1.0.1). At the same time, the KANs were adapted to the target dataset using the Python package pykan (version 0.2.8).

#### 2.6.1 Ensemble Models

The ensemble models employed (ERF, XGB and GBM) were selected for their ability to capture complex relationships in data, offering strong predictive performance and interpretability ^43–45^. ERF extends the traditional Random Forest algorithm by building numerous unpruned decision trees on random subsets of the data, with predictions made by averaging the outputs across the trees. This approach reduces variance and overfitting while effectively capturing nonlinear relationships with minimal parameter tuning ^36^. XGB and GBM, both employing boosting techniques, construct trees sequentially, each tree addressing errors from its predecessors. XGB, an optimized gradient boosting method, is renowned for its speed, accuracy, and scalability. It incorporates features such as parallel computations, regularization to mitigate overfitting, and efficient handling of missing data ^37^. Traditional GBM, although less computationally efficient, utilizes gradients to build trees iteratively and adjusts each tree’s impact with a learning rate, ensuring precision in predictions ^38^. Together, these models leverage complementary strengths, bolstering the robustness and interpretability of BMI predictions.

#### 2.6.2 Linear Models

The three linear models (EN, Lasso, and Ridge regression) were chosen for their effectiveness in handling multicollinearity and their ability to deliver interpretable results in regression tasks. Lasso regression employs an *L*1 penalty, which drives certain coefficients to zero, effectively performing feature selection by retaining only the most significant predictors. This approach simplifies the model and enhances interpretability by reducing the number of included variables ^40^. Ridge regression, in contrast, applies an *L*2 penalty that shrinks all coefficients toward zero without eliminating them ^41^. This regularization technique enhances model stability, particularly when predictors are highly correlated, but does not perform feature selection. The EN combines the strengths of Lasso and Ridge by incorporating both *L*1 and *L*2 penalties, striking a balance between feature selection and coefficient stability. This makes EN particularly effective in scenarios involving strong multicollinearity or when the number of predictors exceeds the number of observations ^39^. The three models offer a robust and adaptable framework for managing complexity and interpretability in regression analysis.

#### 2.6.3 Multi-Layer Perceptron

The Multi-Layer Perceptron (MLP), a type of neural network, excels at modeling complex, nonlinear relationships in regression tasks such as predicting childhood obesity. With multiple layers of nodes and nonlinear activations, MLPs effectively capture intricate patterns ^46^. However, their “black box” nature reduces interpretability, which is addressed by methods like SHAP, LIME, and permutation importance ^12^. Despite requiring large datasets, MLPs offer a powerful framework for complex regression problems.

#### 2.6.4 Kolmogorov-Arnold Network

The KAN, a deep learning model ^16^, is inspired by the Kolmogorov-Arnold Representation Theorem, which states that any multivariate continuous function *f*(*x*_1_, *x*_2_, …, *x*_*n*_) can be decomposed into a sum of univariate functions:

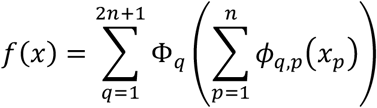

where *ϕ*_*q,p*_(*x*_*p*_) are learnable univariate functions. Unlike traditional neural networks, KANs represent activation functions as spline-based transformations on edges rather than fixed functions on nodes. The general deep KAN model is defined as:

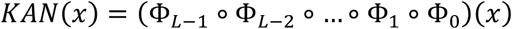

where each layer transforms inputs via learnable splines.

Each edge function in KANs is represented by a B-spline:

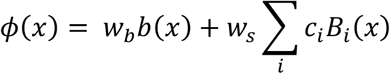

where:

- 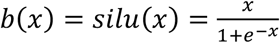 is a residual basis function.
- *B*_*i*_(*x*) are B-spline basis functions with trainable coefficients *c*_*i*_.
- *w*_*b*_ and *w*_*s*_ control activation magnitudes.

The tuning involves grid extension, which defines the B-spline resolution, and regularization of lambda, *L*1 and entropy penalties to induce sparsity. In this study, we optimized the hyperparameters by searching a grid extension between 3 and 5, lambda from 0.01, 0.05, 0.1, and 0.2, *L*1 from 1, 2, and 5, and entropy from 1, 2, and 5.

KANs utilize sparsification and pruning to reduce complexity and improve interpretability by removing less important nodes. The importance score of a node at the *l* -th layer *i* -th node is computed as

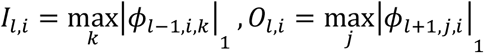

where *k* and *j* are the indices of the nodes at the layer below and above the current layer *l*. Nodes are pruned if:

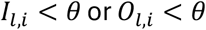

where *θ* is a predefined cut-off threshold. In this study, we pruned the model by setting the cut-off values as 0.01 for nodes and 0.03 for edges. This indicates that if the attribution score of a node or edge is below the cut-off value, it was considered dead and was set to zero. This removes weak connections while maintaining model performance.

After pruning, we further symbolized the expressions by constraining edge functions to known forms from the function library, which includes most typically used functions such as linear, polynomial, reciprocal, root, exponential, logarithmic, trigonometric, hyperbolic, Gaussian, a constant zero function and other activation functions. A learned function *φ*(*x*)can be fitted to a known symbolic function from this library via:

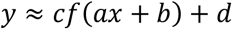

where *a, b, c* and *d* are trainable affine parameters. The final symbolized formula is a compact mathematical expression containing all the input variables *x*_*i*_.

In this study, we demonstrated the use of a shallow KAN model with a single hidden layer between the input and output layers. Consequently, the importance of each input variable is determined by multiplying the activation weights along the paths from the input to the output.

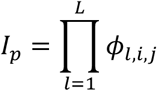

where the importance score *I*_*p*_ reflects how strongly an input influences the output, making KANs more interpretable than MLPs. To facilitate interpretation, the final weights of each input variable were normalized to percentages, representing their relative contributions to the overall model.

### 2.7 Model Training and Evaluation Metrics

All eight models were deployed across Dataset 1, Dataset 2, and their combined datasets 1 and 2. A 5-fold cross-validation strategy with varying randomization was implemented to split the data into training and testing sets. Each model was trained on the training set and evaluated on the testing set, with predictions collected from all five testing folds for further analysis. This approach ensured robust evaluation by mitigating the risk of overfitting and providing reliable performance estimates.

For each model, hyperparameters, including the number of clusters and the *r*_cutoff_ value, were finely tuned. For formalization, we utilized the default formula library to derive formulas for calculating the final predictions. This approach assumes no prior knowledge of the input variables’ distributions or their relationships to the final outcomes.

Model performance was evaluated using three key metrics: the coefficient of determination (R^2^), root mean squared error (RMSE), and Mean Absolute Percentage Error (MAPE), to assess explained variance and prediction accuracy. The best-performing model and dataset were selected based on these metrics for subsequent results presentation and analysis.

- R^2^ (Coefficient of Determination)

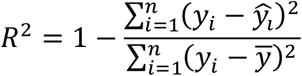

where *y*_*i*_ represents the actual values, 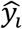 represents the predicted values, and 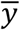 is the mean of the actual values. An R^2^ score closer to 1 indicates better model performance.
- Root Mean Squared Error (RMSE):

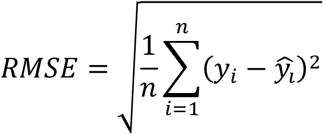
- Mean Absolute Percentage Error (MAPE):

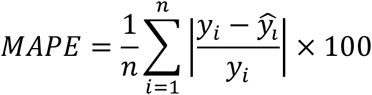

Both RMSE and MAPE closer to 0 represent better performance.

The performance of the KAN models with formalization was compared to that of the original KAN models and the seven conventional models by examining R^2^, RMSE and MAPE. Additionally, the estimates of the KAN models, both with and without formalization, were compared against the true BMI values. We further analyzed key factors and their derived formulas identified by the KAN models, such as year 5 BMI z-scores (Y5BMIz) and the PGS7, for their role in predicting the year 8 BMI.

## 3 Results

### 3.1 Models and Datasets Comparison

Table 4 presents the R^2^, RMSE and MAPE values of eight models trained on Dataset 1, Dataset 2, and their combination. When comparing the impact of datasets on BMI estimation, the combined use of Dataset 1 (early life epidemiology) and Dataset 2 (genetics) yielded the best results, slightly better than using Dataset 1 (early life epidemiology) alone across the eight models. In contrast, using Dataset 2 (genetics) in isolation provided little value in predicting BMI. Among the eight models evaluated, the KAN model generally outperforms the others in terms of predictive accuracy.

**Table 4.** Performance comparison of different models across datasets using R2, RMSE, and MAPE.

| Dataset | Model | $R^2$ | RMSE | MAPE |
| --- | --- | --- | --- | --- |
| 1 | ERF | 0.8 | 1.11 | 4.57 |
|  | XGB | 0.78 | 1.17 | 4.75 |
|  | GBM | 0.79 | 1.14 | 4.71 |
|  | EN | 0.78 | 1.2 | 4.96 |
|  | Lasso | 0.79 | 1.15 | 4.8 |
|  | Ridge | 0.78 | 1.17 | 4.98 |
|  | MLP | 0.77 | 1.19 | 4.94 |
|  | KAN | 0.81 | 1.13 | 4.64 |
| 2 | ERF | 0.12 | 2.38 | 9.74 |
|  | XGB | 0.11 | 2.39 | 9.72 |
|  | GBM | 0.12 | 2.39 | 9.75 |
|  | EN | 0.12 | 2.38 | 9.76 |
|  | Lasso | 0.12 | 2.38 | 9.76 |
|  | Ridge | 0.12 | 2.38 | 9.76 |
|  | MLP | 0.12 | 2.38 | 9.76 |
|  | KAN | 0.12 | 2.39 | 9.75 |
| 1 & 2 | ERF | 0.8 | 1.1 | 4.52 |
|  | XGB | 0.79 | 1.13 | 4.72 |
|  | GBM | 0.81 | 1.07 | 4.53 |
|  | EN | 0.78 | 1.19 | 4.92 |
|  | Lasso | 0.79 | 1.17 | 4.93 |
|  | Ridge | 0.78 | 1.19 | 4.95 |
|  | MLP | 0.77 | 1.18 | 4.93 |
|  | KAN | 0.81 | 1.09 | 4.52 |
|  | KAN-f * | 0.8 | 1.09 | 4.53 |

\* KAN-f represents the formularized KAN model.

These findings highlight that the PGS variables alone (Dataset 2) do not enable the model to achieve strong performance, as the model struggles to identify a clear functional relationship between the PGS variables and BMI. However, when combined with Dataset 1, the correlations between the PGS variables and BMI contribute positively to the training process, enhancing the model’s performance by leveraging the complementary information from both datasets.

### 3.2 Feature Importance

Feature importance for linear and ensemble models was determined using feature coefficients and metrics provided by the scikit-learn 1.7 Python library, leveraging their intrinsic interpretability. Since MLPs lack inherent feature importance attributes due to their complex and nonlinear structure, we employed SHAP, LIME, and permutation importance as ad hoc methods to estimate feature importance for MLPs. The feature importance in the decision-making process of the KAN model was determined by multiplying the weights of each activation function from the leaf nodes (input variables) to the output (year 8 BMI value). Figure 2 presents the aggregated feature importance of selected factors across eight models, evaluated over ten sets. All methods consistently identify the year 5 BMI z-score as the dominant predictor of year 8 BMI, followed by year 5 mid-arm circumference and other anthropometric measurements, such as chest circumference and subscapular skinfold. Linear models assign comparable weights to chest circumference, mid-arm circumference, and subscapular skinfold at year 5. In addition to these anthropometric features, the KAN highlights the mother’s occupation in the first year and the polygenic score (PGS7) as notable predictors.

**Figure 2.**
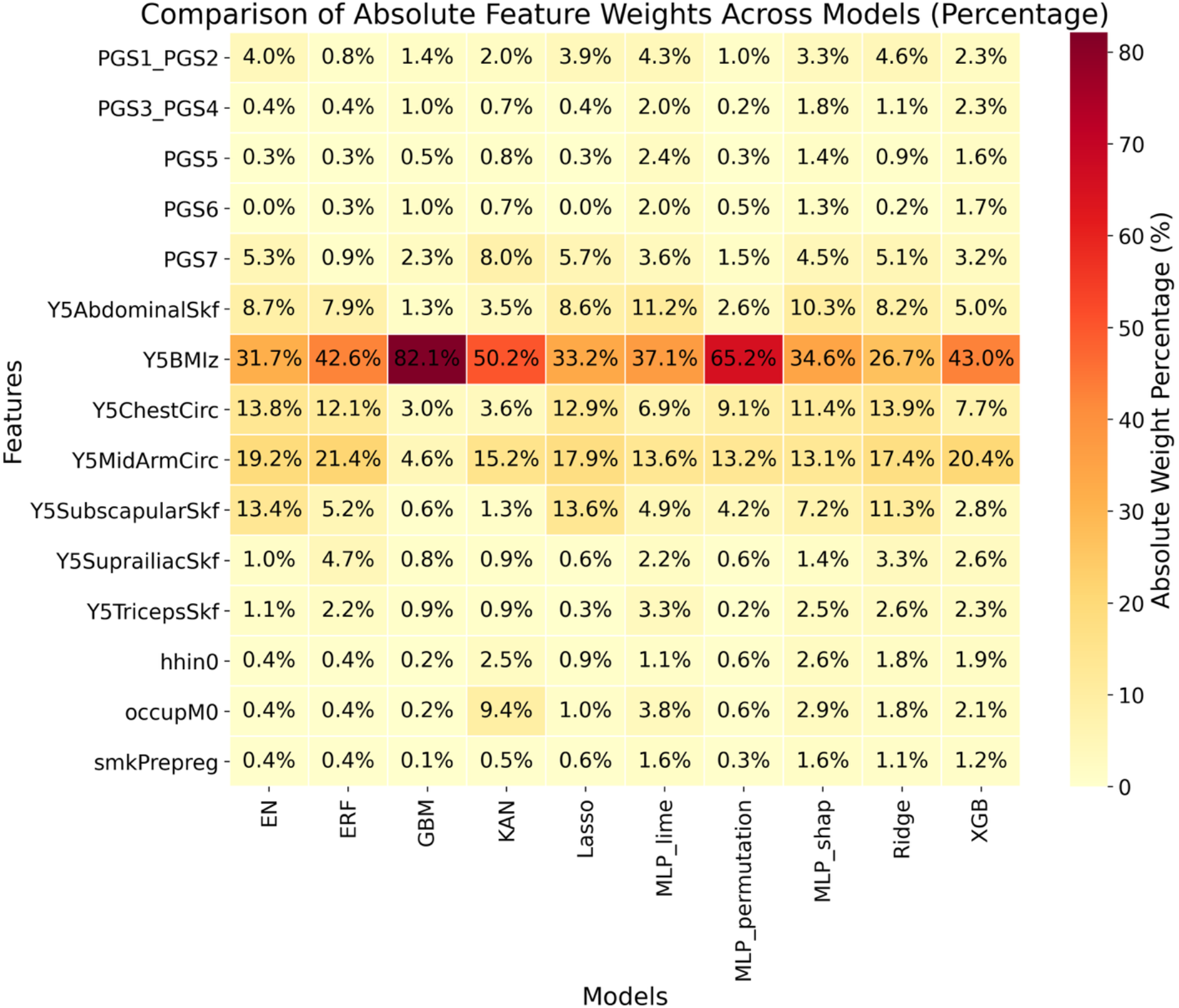
Feature importance across models

Given the superior performance of the KAN compared to other models, we analyzed its decision-making process by examining tree plots before and after pruning. In this study, pruning was performed by setting a node threshold of 0.01 and an edge threshold of 0.03 to simplify the model while preserving its predictive capabilities, as described in 2.6.4. Figure 3 displays the original and pruned tree plots of input variable importance for BMI predictions.

**Figure 3.**
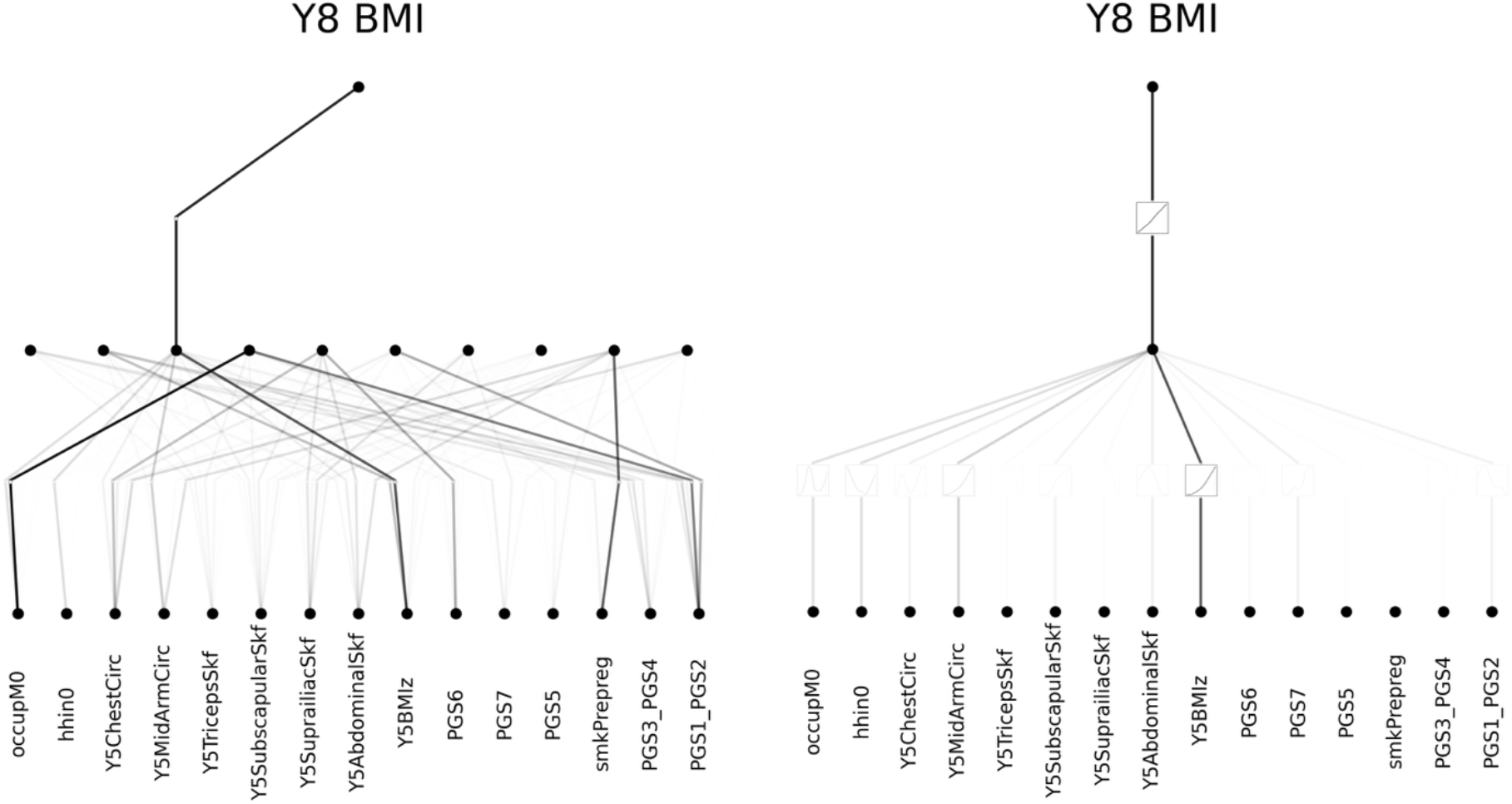
Original (left) and pruned (right) tree plots of KAN. The bottom black points represent the selected factors used in model development, the middle layer points represent the first-layer nodes of the KAN models, and the top point corresponds to the year 8 BMI value. The color intensity of the connecting lines reflects the impact of the lower node on the upper node. For the pruned model, the nail plots in the middle of the connecting lines represent the activation functions used in the models.

### 3.3 Formularization

Beyond analyzing feature importance derived from the model and ad hoc methods such as SHAP, LIME, and permutation importance, the KAN offers a mathematical explanation of its decision-making process. We derived the explicit formulas employed by the KAN model to estimate target outcomes, providing a transparent and interpretable representation of its predictive mechanism. Without making any prior assumptions about the functional form or distribution of the variables, we employed the activation functions recommended by the KAN model package. These functions were chosen as the optimal fit from the available function library, as described in 2.6.4.

We fixed the activation functions and re-evaluated the formularized models on the testing data to obtain the estimated BMI values. Figure 4 compares the performance of the models before and after formularization against the ground truth. The results clearly show that the model maintains comparable performance after formularization. This consistency allowed us to further explore the mathematical functions associated with variables of interest and hypothesize their distributions in predicting target BMI values.

**Figure 4.**
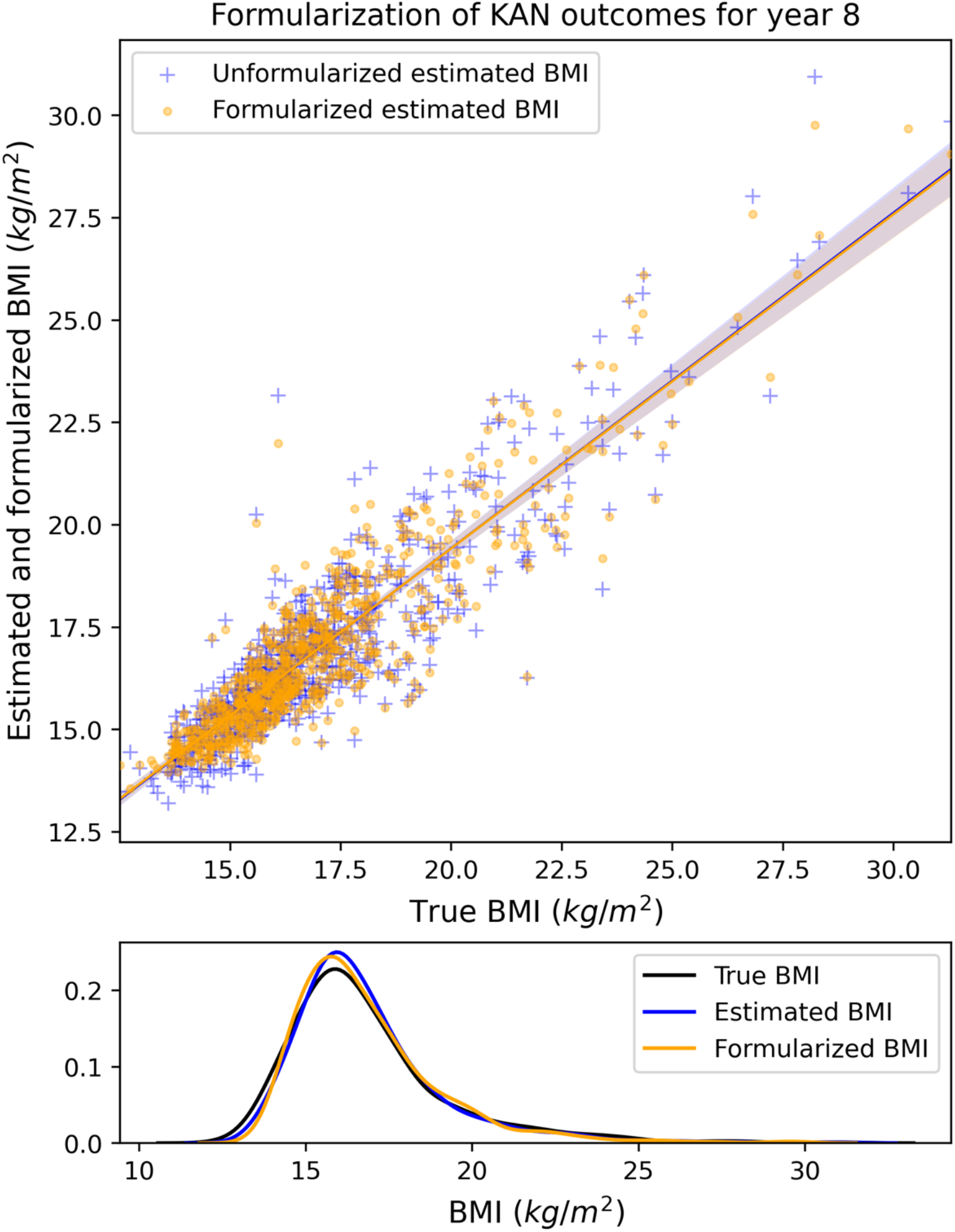
Scatter and distribution plots of the estimated and real BMI values of year 8, before and after formularization

Diving deeper, we analyzed the formulas of the critical features and their relationships with the final outcome, the year 8 BMI. To illustrate the results, we selected Y5BMIz (the BMI z-score at year 5) and PGS7 to demonstrate the mathematical interpretation provided by KAN. The plots for the other important features (Y5MidArmCirc: mid-arm circumference at year 5; and occupM0: occupation of the mother at the birth of the child) are provided in the Supplementary Materials. Our findings reveal that the true and estimated BMI values exhibit an exponential or first-quadrant sine-like functional relationship with Y5BMIz, as shown in Figure 5. In contrast, PGS7 does not follow a distinct functional pattern in relation to the target BMI, as observed in Figure 6. However, a positive correlation is evident between PGS7 and BMI at the age of 8 years.

**Figure 5.**
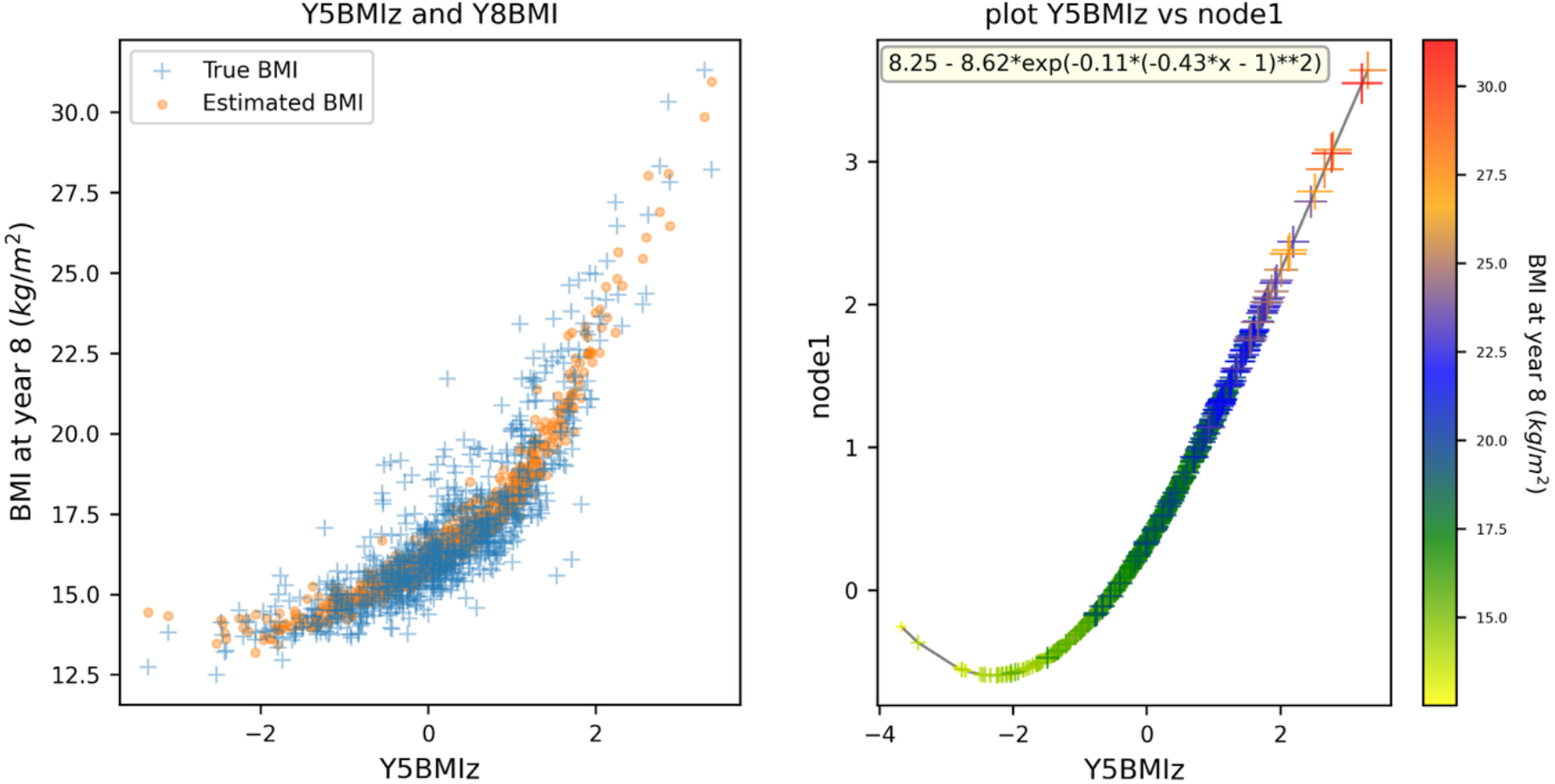
Left - the relationship between Y5BMIz on the x-axis and the true BMI (blue crosses) alongside the model’s estimated BMI values (orange dots) at year 8 on the y-axis. Right - the KAN node function for Y5BMIz, where the data points (crosses) are color-coded and size-scaled to represent BMI values. The functional form after standardization is provided in the top-left corner of the right panel.

**Figure 6.**
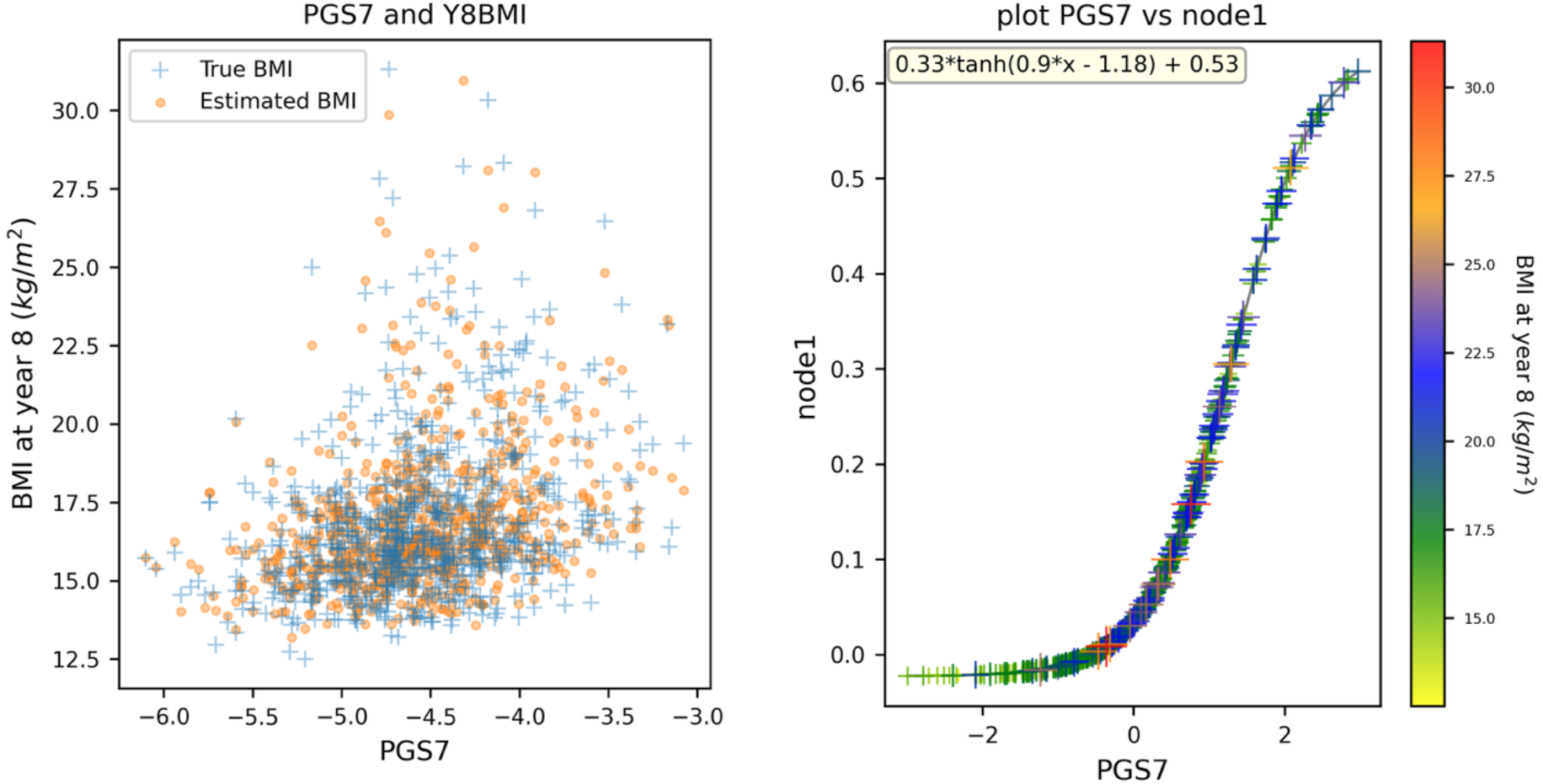
Left - the relationship between PGS7 on the x-axis and the true BMI (blue crosses) alongside the model’s estimated BMI values (orange dots) at year 8 on the y-axis. Right - the KAN node function for PGS7, where the data points (crosses) are color-coded and size-scaled to represent BMI values. The functional form after standardization is provided in the top-left corner of the right panel.

Finally, we aggregated the formularization results, reversed the standardization of the input variables, and derived the overall features formula for predicting BMI at 8 years old, which is:

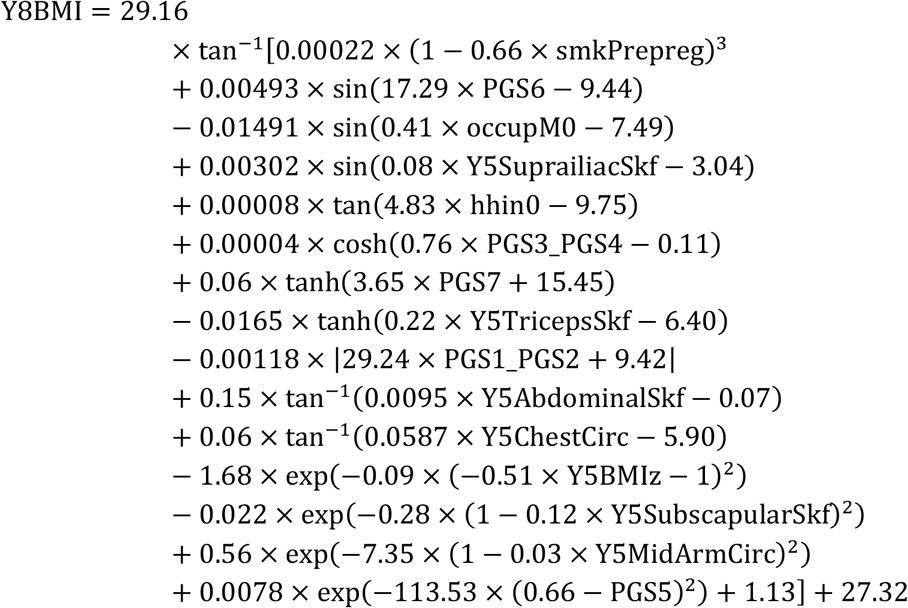

### 3.4 Implementation

To enable practical clinical application, we developed the Year 8 BMI Prediction Calculator, a publicly accessible calculator hosted on the website (https://bmi-y8-calc.onrender.com/), with the original code available on GitHub ^2^. This calculator utilizes mathematically transparent formulas derived from the KAN model, which is trained without PGS for enhanced generalizability. The tool offers two prediction modes: a “Simple” mode using 10 key variables from the KAN-derived formula, including maternal occupation, household income, Year 1 BMI-for-age, and Year 5 anthropometric measurements; and a “Comprehensive” mode employing 20 variables from Dataset 1, incorporating additional factors such as parental heights, gestational age, and early childhood measurements for improved accuracy. Both modes achieve comparable performance (R^2^ = 0.81), enabling flexible and interpretable predictions for clinicians and researchers to support early interventions for obesity. The tool supports individual predictions via an interactive form and batch processing through CSV file uploads, automatically computing derived variables like BMI z-scores. Built with modern web technologies, this application empowers healthcare professionals, researchers, and educators to identify children at risk of weight-related health issues early, enabling timely interventions and personalized health planning.

## 4 Discussion

This study demonstrates that the Kolmogorov-Arnold Network (KAN) model outperforms traditional machine learning models in predicting childhood obesity at age 8, achieving an R^2^ of 0.81 and RMSE of 1.09 when combining epidemiological and genetic data. The KAN’s interpretability revealed that the year 5 BMI z-score is the dominant predictor, followed by mid-arm circumference, maternal occupation, and polygenic score PGS7, offering actionable insights into obesity risk factors.

In biological research, conventional statistical models, such as linear regression or generalized linear models, have long been the cornerstone for analyzing relationships between input variables and outcomes. These models are widely favored for their simplicity, transparency, and ease of implementation. However, they inherently assume linearity in the relationships between input variables, among input variables and between inputs and the target outcome. While such assumptions work well for data with straightforward, linearly distributed relationships, they fail to handle complex datasets. For instance, input variables in biological studies often exhibit intricate interdependencies, nonlinear distributions, and hierarchical relationships, which are difficult to capture with conventional models. Additionally, relying on a single overarching model to fit all variables, irrespective of their distributional properties, may lead to suboptimal performance and inaccurate interpretations.

Deep learning models have emerged as powerful tools to address these limitations. Their capacity to model highly nonlinear and complex relationships has led to significant advancements in various fields, including biology and medicine. However, despite their impressive predictive capabilities, deep learning models often lack interpretability, a critical aspect in fields like biomedical research where understanding the underlying biological mechanisms is as important as achieving high predictive accuracy. The opaque nature of deep learning models has hindered their widespread adoption in scenarios requiring clear and actionable insights.

The KAN model presents a promising alternative to bridge this gap. KAN achieves a balance between predictive performance and interpretability, leveraging its innovative architecture to provide actionable insights. KAN incorporates a formularization mechanism enabled by fixing the activation functions, allowing it to model complex, nonlinear relationships while preserving interpretability. This capability is particularly advantageous for tasks involving heterogeneous input variables with varied distributions and intricate interrelationships.

The results of this study underscore the utility of KAN in BMI prediction. By outperforming traditional models and maintaining a high degree of interpretability, KAN demonstrates its potential as a robust tool for analyzing complex biomedical data. The incorporation of formularization functions provides researchers and clinicians with not only accurate predictions but also valuable insights into the underlying factors driving those predictions.

It is important to acknowledge the limitations of the KAN model. The Kolmogorov-Arnold Representation theorem, on which KAN is based, assumes that any multivariate continuous function on a bounded domain can be expressed as a finite composition of continuous functions of a single variable and the binary operation of addition. This assumption poses challenges when dealing with binary variables, categorical variables with limited bins, or entirely non-continuous discrete variables, as these may not naturally fit into the model’s formulation framework. Future studies should explore strategies to address this limitation, such as integrating piecewise functions into the model architecture. In this study, we observed that certain variables, such as ethnicity and smoking status, are categorical rather than continuous. To enable the model to account for these factors, we applied one-hot encoding where necessary. For categorical variables with ordered bins, such as household income and education levels, we treated them as continuous variables, as the number of bins approximates continuity.

In summary, this study demonstrates the potential of explainable advanced machine learning models, such as KAN, to address and explain the complexities of BMI prediction. By providing both high predictive accuracy and interpretability, KAN offers a potential tool for utilizing the multifaceted influences on BMI in real-world predictions.

## Abbreviations

EN: Elastic Net
ERF: Extreme Random Forest
GBM: Gradient Boosting Machine
KAN: Kolmogorov Arnold Network
Lasso: Least Absolute Shrinkage and Selection Operator
MLP: Multi-Layer Perceptron
PGS: Polygenic Score
RFE: Recursive Feature Elimination
XGB: Extreme Gradient Boosting

## Acknowledgement

We gratefully acknowledge all Raine Study participants and their families for their continued participation in the study, as well as the Raine Study team for their coordination and data collection efforts. We also thank the NHMRC and the Raine Medical Research Foundation for their support. The core management of the Raine Study is funded by The University of Western Australia, Curtin University, The Kids Research Institute Australia, Women and Infants Research Foundation, Edith Cowan University, Murdoch University, The University of Notre Dame Australia and the Western Australian Future Health Research and Innovation Fund (2023-2024; Grant ID WACSOSP2023-2024). The Pawsey Supercomputing Centre provided computation resources to carry out analyses required with funding from the Australian Government and the Government of Western Australia. The data collection of the Raine Study Gen1- and Gen2-1, 2, 5, 8 year follow-ups was funded by NHMRC Grant and The Raine Medical Research Foundation.

## Author Contributions

F.C. designed and conducted the study, developed the model, aggregated and visualized the results, performed the analysis, and drafted the manuscript and supplementary materials. R.H. was responsible for data acquisition, results interpretation, analysis, and manuscript writing. P.M. contributed to data acquisition, model evaluation, and results interpretation and analysis. All authors reviewed and approved the final manuscript.

## Data Availability

The datasets generated and analyzed during this study are not available. The Raine study is committed to a high level of confidentiality of the data in line with the informed consent provided by participants. Requests for data should be directed to the Raine Study Executive.

## Declaration of Generative AI and AI-assisted technologies in the writing process

During the writing process of this paper, the authors used ChatGPT in order to improve language only. After using this tool, the authors reviewed and edited the content as needed and take full responsibility for the content of the publication.

## Appendix

### A. Algorithm for variable clustering and selection

To optimize data cleansing while preserving as many entries as possible, we developed a variable clustering and selection algorithm, applied after Step 6 as described in Figure 1: “Convert weight and height into BMI z-scores, encode parental race”. This algorithm consists of three main stages: Step A (“Deep Clean”), Step B (“Cluster Variables”), and Step C (“Select Important Clusters”). Detailed descriptions of these steps are provided in Figure 7. The input data for this algorithm corresponds to the dataset obtained from Step 6 in Figure 1.

**Figure 7.**
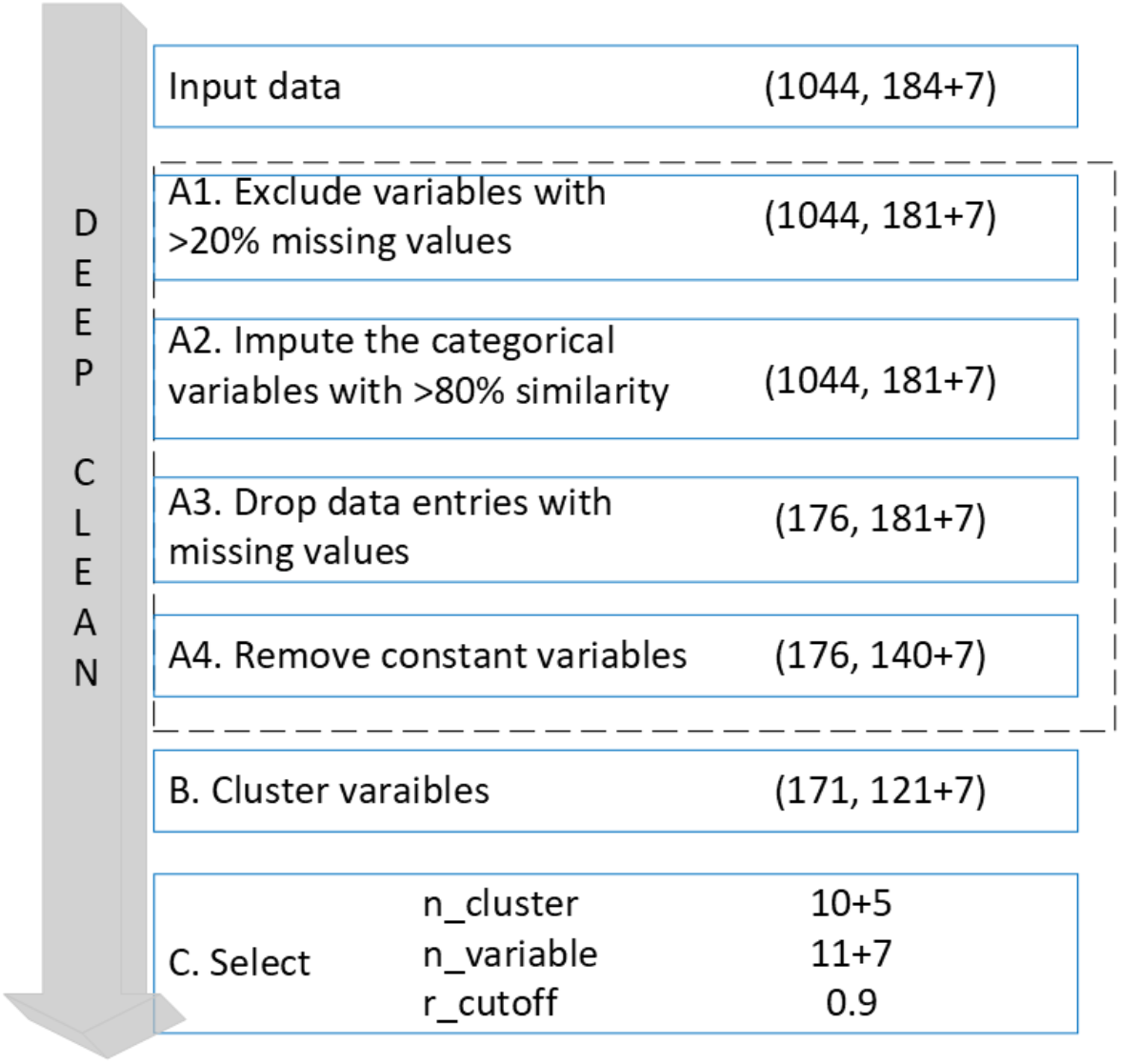
Workflow of variables clustering and selection of Dataset 1 and 2. The form of (i, j+k) represents the number of samples (i), the number of Dataset 1 variables (j) and the number of Dataset 2 PGS (k)

In Step A (“Deep Clean”), we refined the dataset by removing variables with more than 20% missing values, imputing categorical variables with over 80% similarity, and dropping data entries with missing values in the remaining variables. Additionally, constant variables were eliminated. As shown in Figure 7, this process significantly reduced the number of data entries. While such a reduction could adversely impact machine learning model development if used directly for training, we utilized this “clean” data solely for clustering and selection to identify important factors and eliminate redundant variables.

The implementation of Step B (“Cluster Variables”) is illustrated in Figure 8. First, we computed the correlation matrix of the input variables. For any pair of items with a correlation coefficient *r*(*A, B*) exceeding the threshold value *r*_*cutoff*_, we replaced them with a single component X derived through Principal Component Analysis (PCA). Each item could represent either a single variable or a cluster of multiple variables. The *r*_*cutoff*_ value was tuned during model training and selected from options of 0.6, 0.7, 0.8, and 0.9.

**Figure 8.**
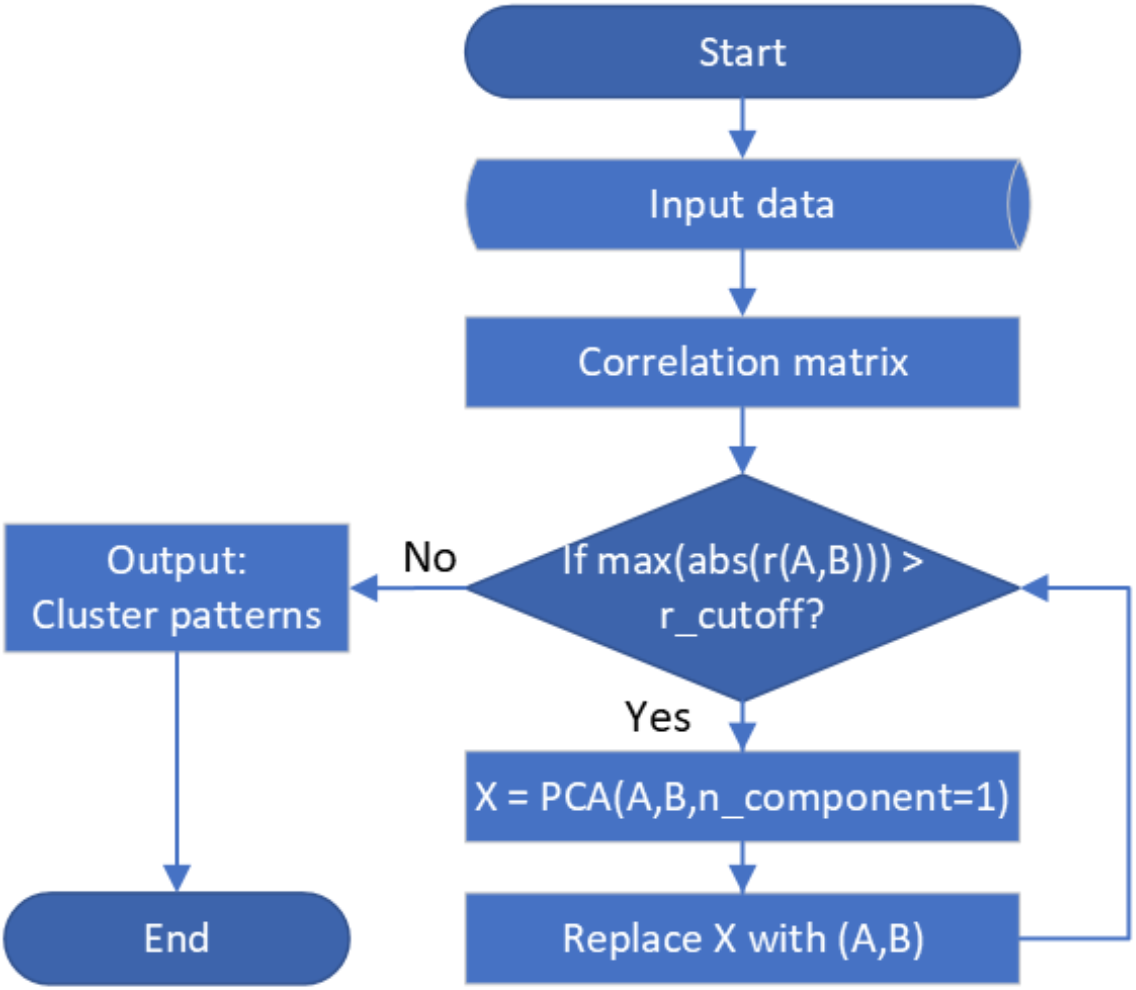
Correlated-based clustering algorithm.

After clustering, we obtained both the cluster patterns and the clustered dataset for Step C (“Select *n*_*cluster*_, *n*_*variable*_, *r*_*cutoff*_”) and subsequent analyses. These clusters were treated as independent units, exhibiting minimal dependency on one another.

In Step C (“Selection”), we employed Recursive Feature Elimination (RFE) integrated with ERF to identify potential predictors. The optimal number of predictors was determined by tuning among 10, 20, 30, and 40.

## Footnotes

1 The variables in the Raine Study are mostly harmonized with the LifeCycle Project-EU Child Cohort Network^21^

2 https://github.com/ICRAR/BMI_y8_calc

## References

1. Rubino F, Cummings DE, Eckel RH, et al. Definition and diagnostic criteria of clinical obesity. Lancet Diabetes Endocrinol. 2025;13(3):221–262. doi:10.1016/S2213-8587(24)00316-4

2. Colmenarejo G. Machine learning models to predict childhood and adolescent obesity: A review. Nutrients. 2020;12(8). doi:10.3390/nu12082466

3. Safaei M, Sundararajan EA, Driss M, Boulila W, Shapi’i A. A systematic literature review on obesity: Understanding the causes & consequences of obesity and reviewing various machine learning approaches used to predict obesity. Comput Biol Med. 2021;136:104754. doi:10.1016/j.compbiomed.2021.104754

4. Gerl MJ, Klose C, Surma MA, et al. Machine learning of human plasma lipidomes for obesity estimation in a large population cohort. PLoS Biol. 2019;17(10):e3000443. doi:10.1371/journal.pbio.3000443

5. Scheinker D, Valencia A, Rodriguez F. Identification of Factors Associated With Variation in US County-Level Obesity Prevalence Rates Using Epidemiologic vs Machine Learning Models. JAMA Netw Open. 2019;2(4):e192884. doi:10.1001/jamanetworkopen.2019.2884

6. Jindal K, Baliyan N, Rana PS. Obesity prediction using ensemble machine learning approaches. In: Sa PK, Bakshi S, Hatzilygeroudis IK, Sahoo MN, eds. Recent Findings in Intelligent Computing Techniques: Proceedings of the 5th ICACNI 2017, Volume 2. Vol 708. Advances in intelligent systems and computing. Springer Singapore; 2018:355–362. doi:10.1007/978-981-10-8636-6_37

7. Montanez CAC, Fergus P, Hussain A, et al. Machine learning approaches for the prediction of obesity using publicly available genetic profiles. In: 2017 International Joint Conference on Neural Networks (IJCNN). IEEE; 2017:2743–2750. doi:10.1109/IJCNN.2017.7966194

8. Pang X, Forrest CB, Le-Scherban F, Masino AJ. Understanding early childhood obesity via interpretation of machine learning model predictions. In: 2019 18th IEEE International Conference On Machine Learning And Applications (ICMLA). IEEE; 2019:1438–1443. doi:10.1109/ICMLA.2019.00235

9. Singh B, Tawfik H. A machine learning approach for predicting weight gain risks in young adults. In: 2019 10th International Conference on Dependable Systems, Services and Technologies (DESSERT). IEEE; 2019:231–234. doi:10.1109/DESSERT.2019.8770016

10. Fergus P, Hussain A, Hearty J, et al. A machine learning approach to measure and monitor physical activity in children to help fight overweight and obesity. In: Huang D-S, Jo K-H, Hussain A, eds. Intelligent Computing Theories and Methodologies: 11th International Conference, ICIC 2015, Fuzhou, China, August 20-23, 2015, Proceedings, Part II. Vol 9226. Lecture notes in computer science. Springer International Publishing; 2015:676–688. doi:10.1007/978-3-319-22186-1_67

11. Gupta M, Phan T-LT, Bunnell HT, Beheshti R. Obesity Prediction with EHR Data: A deep learning approach with interpretable elements. ACM Trans Comput Healthcare. 2022;3(3). doi:10.1145/3506719

12. Lundberg SM, Lee SI. A unified approach to interpreting model predictions. Advances in neural information …. 2017.

13. Ribeiro M, Singh S, Guestrin C. “why should I trust you?”: explaining the predictions of any classifier. In: Proceedings of the 2016 Conference of the North American Chapter of the Association for Computational Linguistics: Demonstrations. Association for Computational Linguistics; 2016:97–101. doi:10.18653/v1/N16-3020

14. Breiman L. Random Forests. Springer Science and Business Media LLC. 2001;45:5– 32. doi:10.1023/a:1010933404324

15. Bajorath J. From scientific theory to duality of predictive artificial intelligence models. Cell Reports Physical Science. 2025;6(4):102516. doi:10.1016/j.xcrp.2025.102516

16. Liu Z, Wang Y, Vaidya S, Ruehle F, Halverson J. Kan: Kolmogorov-arnold networks. arXiv preprint arXiv …. 2024.

17. He C, Ma M, Wang P. Extract interpretability-accuracy balanced rules from artificial neural networks: A review. Neurocomputing. 2020;387:346–358. doi:10.1016/j.neucom.2020.01.036

18. Fan F-L, Xiong J, Li M, Wang G. On interpretability of artificial neural networks: A survey. IEEE Trans Radiat Plasma Med Sci. 2021;5(6):741–760. doi:10.1109/trpms.2021.3066428

19. Simmonds M, Llewellyn A, Owen CG, Woolacott N. Predicting adult obesity from childhood obesity: a systematic review and meta-analysis. Obes Rev. 2016;17(2):95–107. doi:10.1111/obr.12334

20. Straker L, Mountain J, Jacques A, et al. Cohort Profile: The Western Australian Pregnancy Cohort (Raine) Study-Generation 2. Int J Epidemiol. 2017;46(5):1384–1385j. doi:10.1093/ije/dyw308

21. Jaddoe VWV, Felix JF, Andersen A-MN, et al. The LifeCycle Project-EU Child Cohort Network: a federated analysis infrastructure and harmonized data of more than 250,000 children and parents. Eur J Epidemiol. 2020;35(7):709–724. doi:10.1007/s10654-020-00662-z

22. Ibrahim JG, Molenberghs G. Missing data methods in longitudinal studies: a review. Test (Madr). 2009;18(1):1–43. doi:10.1007/s11749-009-0138-x

23. Wei R, Ogden CL, Parsons VL, Freedman DS, Hales CM. A method for calculating BMI z-scores and percentiles above the 95th percentile of the CDC growth charts. Ann Hum Biol. 2020;47(6):514–521. doi:10.1080/03014460.2020.1808065

24. Organization WH. WHO Child Growth Standards: Length/Height-for-Age, Weight-for-Age, Weight-for-Length, Weight-for-Height and Body Mass Index-for-Age: Methods and …. apps.who.int; 2006.

25. Min J, Wen X, Xue H, Wang Y. Ethnic disparities in childhood BMI trajectories and obesity and potential causes among 29,250 US children: Findings from the Early Childhood Longitudinal Study-Birth and Kindergarten Cohorts. Int J Obes (Lond). 2018;42(9):1661–1670. doi:10.1038/s41366-018-0091-4

26. Guerrero AD, Mao C, Fuller B, Bridges M, Franke T, Kuo AA. Racial and ethnic disparities in early childhood obesity: growth trajectories in body mass index. J Racial Ethn Health Disparities. 2016;3(1):129–137. doi:10.1007/s40615-015-0122-y

27. Guyon I, Elisseeff A. An introduction to variable and feature selection. Journal of machine learning research. 2003.

28. Lambert SA, Wingfield B, Gibson JT, et al. Enhancing the Polygenic Score Catalog with tools for score calculation and ancestry normalization. Nat Genet. 2024;56(10):1989–1994. doi:10.1038/s41588-024-01937-x

29. Weissbrod O, Kanai M, Shi H, et al. Leveraging fine-mapping and multipopulation training data to improve cross-population polygenic risk scores. Nat Genet. 2022;54(4):450–458. doi:10.1038/s41588-022-01036-9

30. Privé F, Aschard H, Carmi S, et al. Portability of 245 polygenic scores when derived from the UK Biobank and applied to 9 ancestry groups from the same cohort. Am J Hum Genet. 2022;109(1):12–23. doi:10.1016/j.ajhg.2021.11.008

31. Khera AV, Chaffin M, Wade KH, et al. Polygenic Prediction of Weight and Obesity Trajectories from Birth to Adulthood. Cell. 2019;177(3):587-596.e9. doi:10.1016/j.cell.2019.03.028

32. Monti R, Eick L, Hudjashov G, et al. Evaluation of polygenic scoring methods in five biobanks shows larger variation between biobanks than methods and finds benefits of ensemble learning. Am J Hum Genet. 2024;111(7):1431–1447. doi:10.1016/j.ajhg.2024.06.003

33. Shim I, Kuwahara H, Chen N, et al. Clinical utility of polygenic scores for cardiometabolic disease in Arabs. Nat Commun. 2023;14(1):6535. doi:10.1038/s41467-023-41985-1

34. Ma Y, Patil S, Zhou X, Mukherjee B, Fritsche LG. ExPRSweb: An online repository with polygenic risk scores for common health-related exposures. Am J Hum Genet. 2022;109(10):1742–1760. doi:10.1016/j.ajhg.2022.09.001

35. Borisevich D, Schnurr TM, Engelbrechtsen L, et al. Non-linear interaction between physical activity and polygenic risk score of body mass index in Danish and Russian populations. PLoS ONE. 2021;16(10):e0258748. doi:10.1371/journal.pone.0258748

36. Geurts P, Ernst D, Wehenkel L. Extremely randomized trees. Mach Learn. 2006;63(1):3–42. doi:10.1007/s10994-006-6226-1

37. Chen T, Guestrin C. XGBoost: A Scalable Tree Boosting System. In: Proceedings of the 22nd ACM SIGKDD International Conference on Knowledge Discovery and Data Mining -KDD ‘16. ACM Press; 2016:785–794. doi:10.1145/2939672.2939785

38. Friedman JH. Greedy function approximation: a gradient boosting machine. Ann Statist. 2001;29(5):1189–1232. doi:10.1214/aos/1013203451

39. Zou H, Hastie T. Regularization and variable selection via the elastic net. J Royal Statistical Soc B. 2005;67(2):301–320. doi:10.1111/j.1467-9868.2005.00503.x

40. Tibshirani R. Regression shrinkage and selection via the lasso. Journal of the Royal Statistical Society: Series B (Methodological). 1996;58(1):267–288. doi:10.1111/j.2517-6161.1996.tb02080.x

41. Hoerl AE, Kennard RW. Ridge Regression: Biased Estimation for Nonorthogonal Problems. Technometrics. 1970;12(1):55–67. doi:10.1080/00401706.1970.10488634

42. Rumelhart DE, Hinton GE, Williams RJ. Learning representations by back-propagating errors. Nature. 1986;323(6088):533–536. doi:10.1038/323533a0

43. Konstantinov AV, Utkin LV. Interpretable machine learning with an ensemble of gradient boosting machines. Knowledge-Based Systems. 2021;222:106993. doi:10.1016/j.knosys.2021.106993

44. Palkar A, Dias CC, Chadaga K, Sampathila N. Empowering glioma prognosis with transparent machine learning and interpretative insights using explainable AI. IEEE Access. 2024;12:31697–31718. doi:10.1109/ACCESS.2024.3370238

45. Du H, Yang Q, Ge A, Zhao C, Ma Y, Wang S. Explainable machine learning models for early gastric cancer diagnosis. Sci Rep. 2024;14(1):17457. doi:10.1038/s41598-024-67892-z

46. Haykin S. Neural Networks: A Comprehensive Foundation. dl.acm.org; 1994.

