## supplementary for "Explainable AI to predict a complex multifactorial outcome, childhood obesity: Application to clinical epidemiology"

**Supplementary – Formularization of the important factors in estimating BMI at 8 years**

Left - the relationship between the factor on the x-axis and the true BMI (blue crosses) alongside the model's estimated BMI values (orange dots) at year 8 on the y-axis. Right - the KAN node function for the factor, where the data points (crosses) are color-coded and size-scaled to represent BMI values. The functional form after standardization is provided in the top-left corner of the right panel.

| 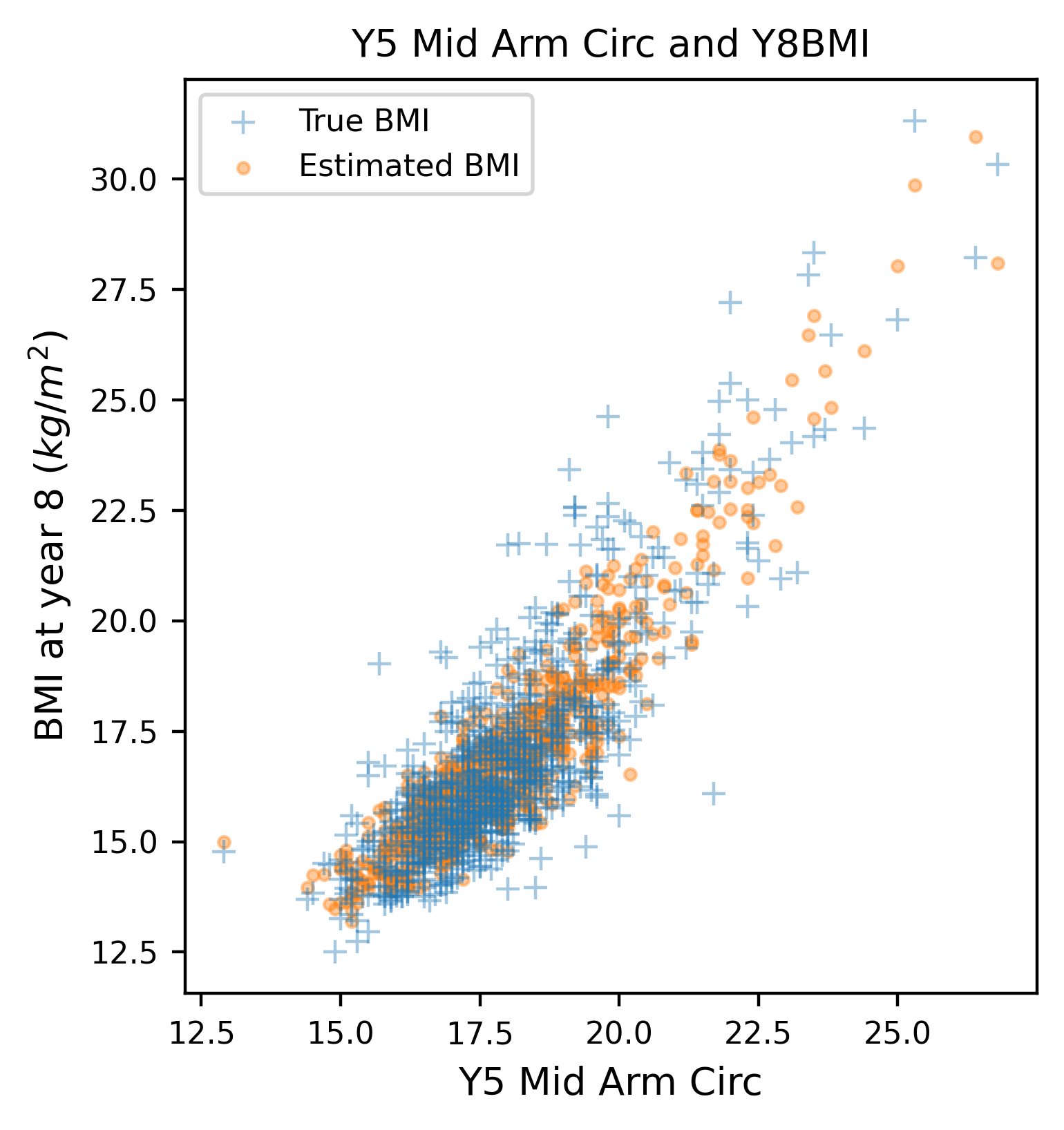 | 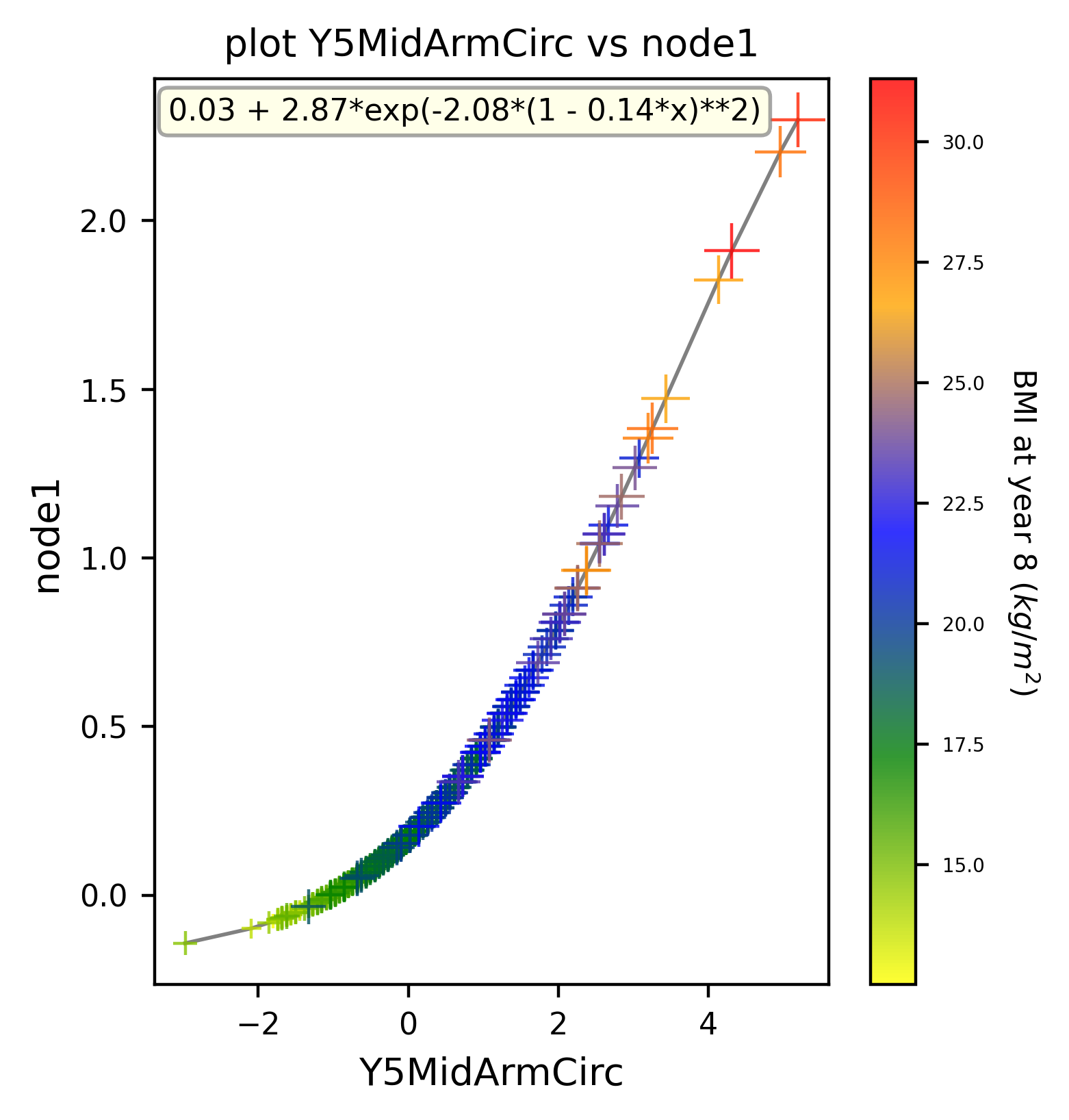 |
| --- | --- |
| 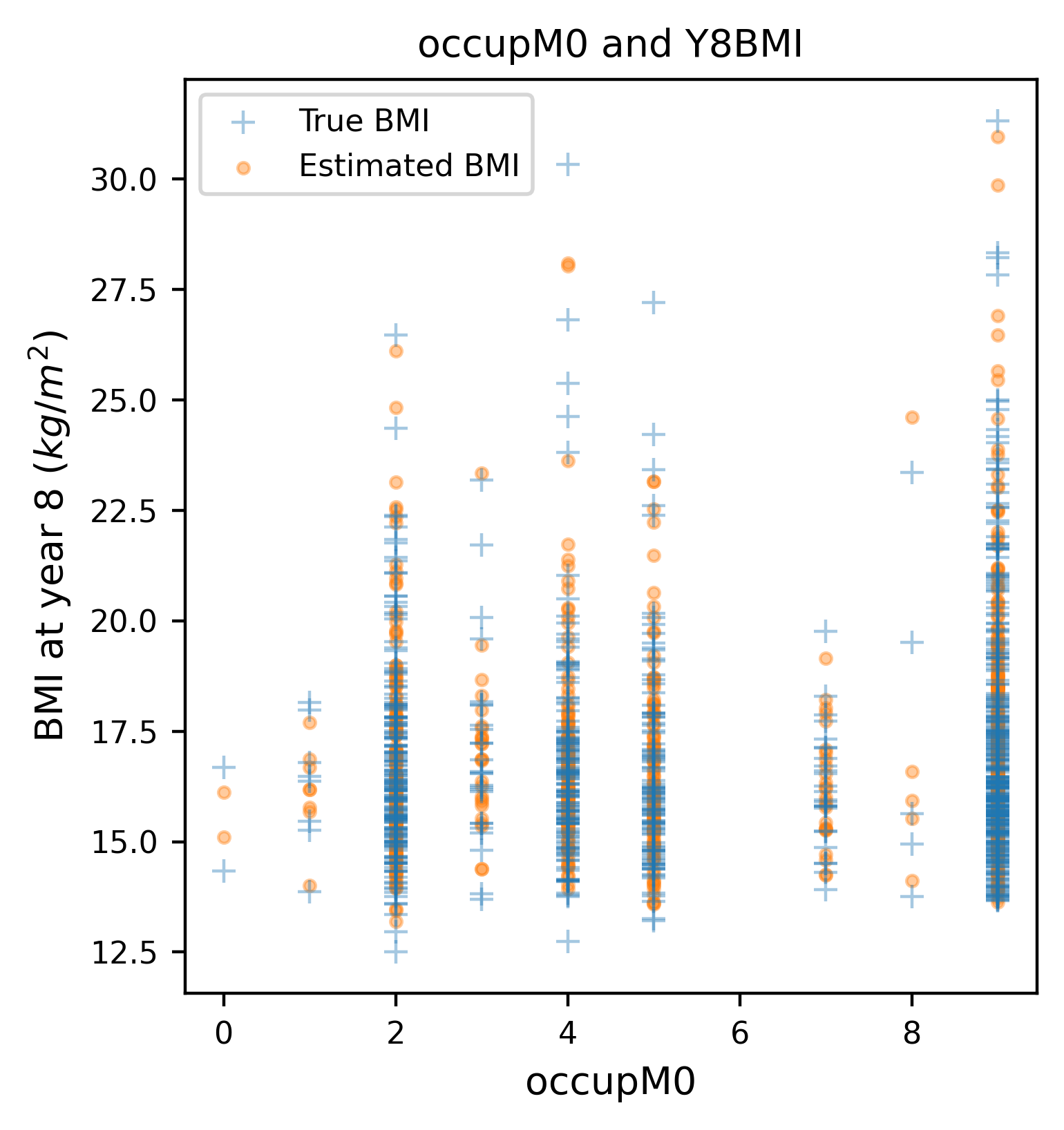 | 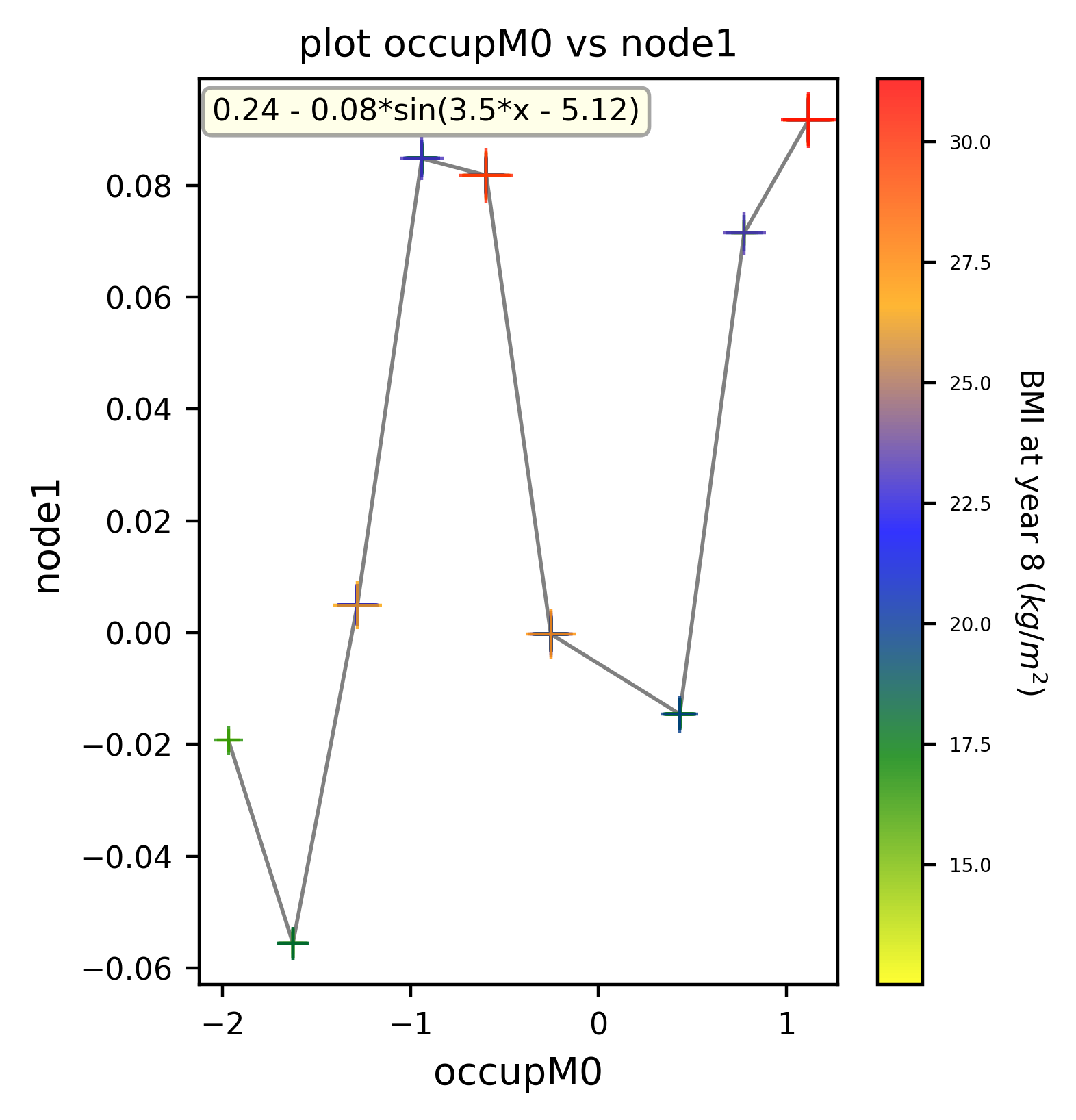 |
